# Integrative Multi-omics of Gynecological Tumors Identifies Novel Singular Biomarkers of Disease Progression

**DOI:** 10.1101/2025.02.21.25322653

**Authors:** Sangeetha Muthamilsevan, Ashok Palaniappan

## Abstract

Gynecological cancers represent a cluster of largely preventable and treatable diseases afflicting women, but with persistent substantial global burden of disease complicated by extant social factors especially in developing countries. Cervical, ovarian and endometrial cancers comprise the major gynecological cancers that might benefit from early-stage diagnosis and personalized treatment strategies. In this work, we performed integrative multi-omics analysis of public-domain omics datasets from The Cancer Genome Atlas consortium and coupled it with custom protocols to identify consensus candidate biomarkers of each of the major gynecological cancers. Such consensus biomarkers were individually evaluated for their ability to classify ‘cancer’ and ‘normal’ samples and those with AUROC > 0.9 were identified as singular biomarkers. Our study yielded the following singular biomarkers: (i) endometrial cancer: MYOZ2, CYP1B1, PPP1R3C, DNASE1L3, ADAMTSL1, LRCH2, RBM20, LOC284276, FAM78B, COL14A1, and PDZRN3; (ii) ovarian cancer: C7 and LONRF2; and (iii) cervical cancer: HAND2, C1QTNF7, JAM3 (with AUROC > 0.99) as well as HSPB7, ACTA2 and DACT7. We demonstrated that factors from multi-omics analysis of endometrial cancer enabled a geometric separation of the ‘cancer’ and ‘normal’ samples. Our results could encourage further research into the multi-omics – based subtypes of these cancers. Our methods could be extended to the analysis of datasets of other cancer types and our studies could pave the way for the development of integrated screening models for the major gynecological cancers.

## INTRODUCTION

Gynecological cancers represent a group of malignancies affecting the female reproductive system, with significant impact on women’s health and ripple effects on family welfare. The major gynecological cancers include the cervical, ovarian, and uterine (chiefly the endometrial) cancers, each with distinct and heterogeneous manifestations, risk factors, and treatment approaches. Cervical cancer remains one of the most common gynecological malignancies worldwide, with an estimated 662,301 cases and 348,874 deaths annually, especially in low-and middle-income countries [1]. The Global Strategy to Accelerate the Elimination of Cervical Cancer (launched in 2020) set an ambitious “90-70-90” target for 2030 [2]. Ovarian cancer carries the highest mortality rate among gynecological cancers in developed nations, with a five-year survival rate around 45% due to late-stage diagnosis in approximately 75% of cases [3]. High-grade serous carcinoma remains the most lethal subtype of ovarian cancer, accounting for approximately 70% of cases [4]. Endometrial cancer mainly affects post-menopausal women, but obesity, metabolic syndrome, and hyperestrogenism are associated with increased risk in younger women [5,6]. Endometrial cancers have been classically classified into four histological types based on the expression of various markers: low-grade endometrioid, high-grade endometrioid, serous, and clear cell EC [8]. Given that histopathological findings in high-grade endometrial cancers could be prone to misdiagnosis [8,9], the molecular classification of endometrial cancers into four distinct subtypes by The Cancer Genome Atlas (TCGA) [10] has proved more useful in understanding disease progression and treatment selection [11]. It has also led to the development of the ProMisE molecular classification system that uses a combination of immunohistochemistry and mutation analysis to divide cases into four prognostic groups: MMRd, POLE, p53abn, and NSMP [12,13].

Research priorities for gynecological cancers include the development of early detection methods (including blood-based technologies), prognostic stratification based on molecular subtype (for e.g. [14]), and identification of novel therapeutic targets toward achieving personalized medicine approaches. The latter includes emerging treatment options such as the optimization of immunotherapy combinations. Studies of resistance mechanisms are crucial to treatment management. Effective strategies for implementing risk-reduction are vital to the prevention of these cancers.

In this setting, the exploration of multi-omics strategies has revolutionized cancer research by providing a holistic view of disease necessary to untangle inherent complexity and heterogeneity. High-dimensional data obtained from high-throughput technologies at multiple levels has propelled a data-driven paradigm shift in medicine [15]. Multimodal integration of perspectives reveals consensus biomarkers and intrinsic patient clusters invisible to single-omics modalities [16]. It is a critical step to acquire knowledge of the unknown void in cancer physiology toward effective individualized diagnosis, targeted therapy, and support for clinical decision-making [17–19]. Different computational strategies are needed to identify biomarkers from different types of omics data [20,21]. In this work, we have used multi-omics approaches and TCGA datasets to identify candidate novel biomarkers for the major gynecological cancers, namely of the ovary, uterus, and cervix. We have performed a custom integration of omics modalities that affect transcript abundance, namely: RNA-Seq, methylomics, miRNA-Seq, and copy number variation.

## MATERIALS AND METHODS

### Datasets

For all the major gynecological cancers, multi-omics profiles were downloaded from the firebrowse.org portal [10]. The corresponding clinical metadata was also retrieved from firebrowse.org and used to annotate the stage information (encoded in the ‘patient.stage_event.clinical_stage’ variable) of the tumor samples and then merged with the omics datasets [10]. In the case of ovarian cancer, GTEx samples with the site as ‘ovary’ were used to augment the number of control samples [22].

### Stage-salient gene identification

A custom biomarker analysis of each omics dataset followed the bi-level protocol developed in our lab [23, 24, 25]; in summary:

(i) Level-I: The gene features specific to a clinical variable were identified using a two-tier contrast. A contrast between the controls on the one hand and each class in the clinical variable of interest on the other could identify class-specific gene biomarkers. In the case of AJCC staging classes, this contrast is performed using the following design matrix:

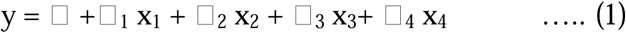

where the independent variables (x_i_) indicate the class of the clinical variable of interest, here Stage I, Stage II, Stage III, and Stage IV. The intercept α is the baseline expression estimated from the controls, and β_i_ represents the log fold change (lfc) in the mean expression of the respective stages (i) with respect to the controls.

(ii) Level-II: Contrast between the various classes of the clinical variable of interest could identify class-salient biomarkers. This between-class contrast is performed for all pairs of classes, using the following design matrix:

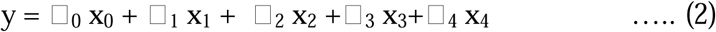

where the controls themselves constitute one of the indicator variables (x_0_), and the β_i_ are coefficients estimated from only the corresponding samples.

By supplying the contrast matrix as an argument to the `contrastsFit` in limma [26], we obtained the log fold change and associated P-value for each contrast in both level-I and level-II. The contrasts against controls were subjected to a stronger significance threshold (adj. P <.001) than that for the between-stages pairwise contrasts (P <.05). In addition, an |lfc| threshold > 2 was uniformly used, constituting constraints that yielded significantly overexpressed or downregulated stage-specific genes (level-I) and then stage-salient genes (level-II).

### Integrative multi-omics analysis

Two major techniques for the analysis and integration of multi-omics datasets were explored in this study, namely Multi-omics factor analysis (MOFA) [27] and Multi-omics integration and Visualization In Cancer Subtyping (MOVICS) [28]. MOFA represents a statistical technique for unsupervised integration of many omics data modalities based on a generalized principal component analysis (PCA). MOFA derives an interpretable low-dimensional representation of data relating to (hidden) factors given several omics data matrices. The determination of cellular states or disease subgroups is facilitated by the identification of primary sources of variation across data modalities. To establish connections between the various –omics, it is crucial that MOFA determines whether the root cause of heterogeneity is specific to one data modality or appears in numerous modalities. MOVICS, on the other hand, integrates ten clustering algorithms to generate consensus features that could help subtype the cancer. Feature selection techniques are provided to obtain reduced gene feature sets from the integrative analysis of RNA-Seq, miRNA-Seq, and DNA methylation multi-omics modalities. Such techniques include Standard deviation (SD, top 100 features), Median absolute deviation (MAD, top 10% of features), PCA (based on threshold variance explained), and univariate Cox analysis (p-value < 0.05). We also used other integrative multivariate techniques like regularized Canonical Correlation Analysis (rCCA) and sparse Partial Least Squares (sPLS) to measure the correlation between the different datasets. Regularization (with L_2_ or ridge regression penalty) facilitates CCA in high dimensions [29]. Similarly, sPLS is an adaptation of the standard PLS technique to apply L_1_ (or lasso regression) based dimension reduction in high-dimensional datasets [30]. The biomarkers identified from integrative multi-omics analysis and the candidate stage-salient biomarkers identified from independent omics analysis were filtered to yield a consensus. The consensus biomarkers were individually evaluated for their ability to discriminate between cancer and control samples using Receiver Operating Characteristic (ROC) curve analysis, and those with area under ROC curve > 0.9 were identified as singular biomarkers of the respective cancer.

## RESULTS

Table 1 shows the stage-wise count of samples with genome-wide omics profiles in all four omics modalities of interest obtained from TCGA. The RNA-Seq profiles comprised 20,501 genes, miRNA-Seq profiles contained 2588 miRNAs, and methylation profiles included ∼480,000 CpG sites (450K arrays). Data preprocessing of the omics profiles was performed per respective published workflows [23–25] and log_2_ transformation of GTEx ‘ovary’ controls was executed. Given the fewer control samples in CESC, it was sought to integrate the RNA-Seq and miRNA-Seq profiles in a bi-omics analysis.

**Table 5.1:** Stage-wise distribution of TCGA multi-omics samples of the major gynecological cancers. For UCEC and CESC, counts of complete profiles (i.e, availability of omics profiles in all modalities of interest, namely RNA-Seq, miRNA, methylation & copy-number variations) are provided. In the case of OV, no controls were available in TCGA necessitating augmentation with GTEx data. Further only one OV sample (with RNA-Seq profile) was available for stage-I cancer, which was then dropped from the analysis.

| Label | OV | UCEC | CESC |
| --- | --- | --- | --- |
| Control |  | 23 | 3 |
| Stage 1 |  | 96 | 163 |
| Stage 2 | 18 | 22 | 70 |
| Stage 3 | 229 | 45 | 46 |
| Stage 4 | 37 | 9 | 21 |

Applying the two-level contrast for the mRNA expression data of each of the cancers yielded the stage-salient genes, whose counts are shown in Table 2. The top ten stage-salient genes of each cancer are identified in Table 3.

**Table 2:**
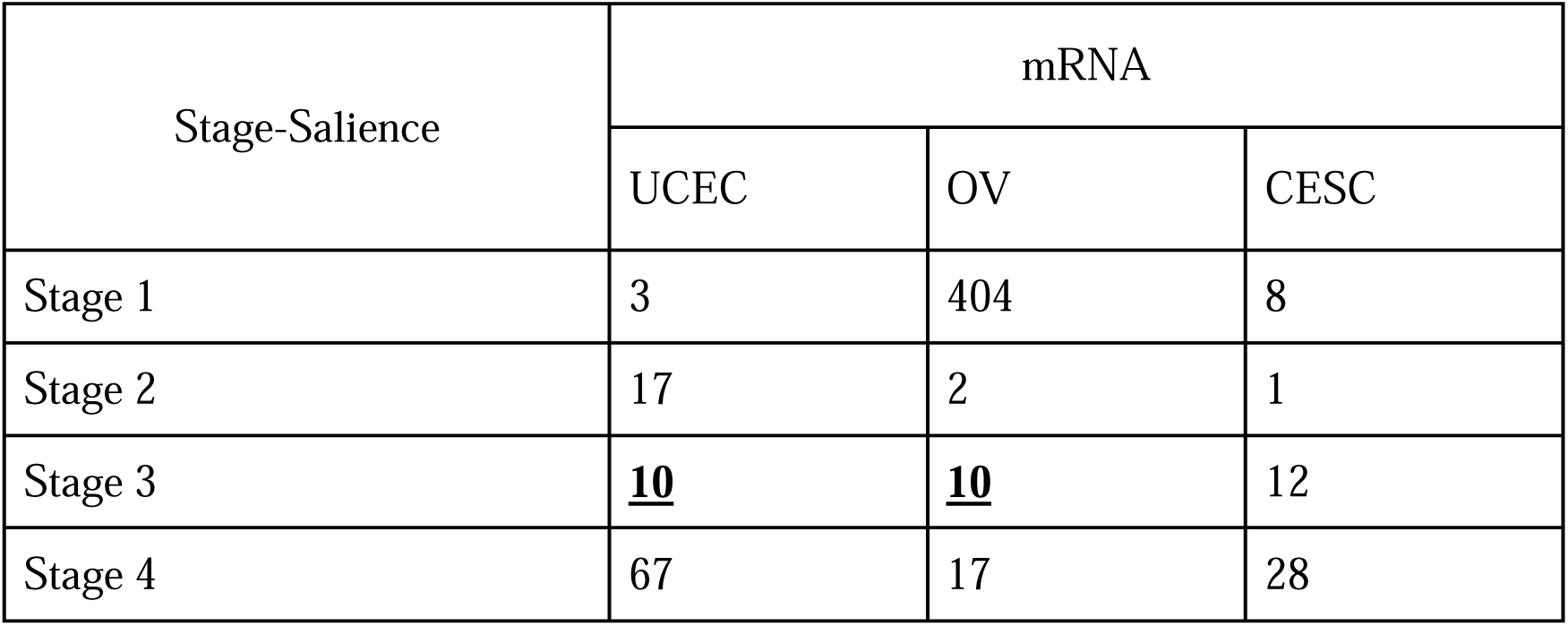
Distribution of cancer stage-salient genes identified in each cancer of interest based on gene expression RNAseq data. An abundance of markers with respect to stage-I ovarian cancer may be explained by the sample size for that stage. If the filters produced no salient genes for a stage, the pval_product quantity was used to identify top ten biomarkers (emphasized).

| Stage-Salience | mRNA |  |  |
| --- | --- | --- | --- |
|  | UCEC | OV | CESC |
| Stage 1 | 3 | 404 | 8 |
| Stage 2 | 17 | 2 | 1 |
| Stage 3 | **<u>10</u>** | **<u>10</u>** | 12 |
| Stage 4 | 67 | 17 | 28 |

**Table 3:** The top stage-salient genes ranked by significance for the cancers of interest. Consensus between the stage-salient genes and selected gene features from the multi-omics analysis are highlighted.

| UCEC |  |  |  |
| --- | --- | --- | --- |
| Stage I | Stage II | Stage III | Stage IV |
| **PPP1R3C** | **KLK12** | **CLDN19** | MAGEA4 |
| **LRCH2** | CLCA2 | MYOD1 | ZNF238 |
| **CYP1B1** | SPINK7 | BTBD17 | CHST13 |
|  | AKR1B10 | **PNOC** | SULT1E1 |
|  | AKR1B15 | KCNK9 | **MAGEA9B** |
|  | RTP1 | SOX11 | SORBS2 |
|  | SPRR3 | BIRC7 | CD1E |
|  | DLX6AS | **RSPO4** | **LOC284276** |
|  | TNFSF11 | **SLC39A5** | CHCHD10 |
|  | KRT16 | HIST1H3G | NXF5 |
| **OV** |  |  |  |
| **Stage I** | **Stage II** | **Stage III** | **Stage IV** |
| **LYSMD3** | MYL3 | RUFY3 | S100A11 |
| **MBLAC2** | ACTC1 | SMG1P7 | RGS10 |
| **CETN3** |  | ZNF594 | LINC01133 |
| MAGEA12 |  | FGF22 | MPZL2 |
| **NOTCH2** |  | RNA5-8S5 | GLDC |
| **GNG12** |  | NKTR | SLC25A3P1 |
| **AUTS2** |  | SLC9A5 | DGKD |
| **AGPAT3** |  | RMRP | C7orf73 |
| **SH2D4A** |  | SNORA73A | ZNF727 |
| **LONRF2** |  | RGPD8 | PDZK1IP1 |
| **CESC** |  |  |  |
| **Stage I** | **Stage II** | **Stage III** | **Stage IV** |
| HBA2 | GPIHBP1 | CLDN8 | LINC02603 |
| HBA1 |  | NMB | PWWP3B |
| HBB |  | MYO15A | CBX8 |
| BCL2L14 |  | ARMCX2 | SMYD2 |
| AGR2 |  | HMSD | RGS13 |
| VENTX |  | MAN1A1 | **HAND2** |
| KRT7 |  | TM4SF1 | HPSE2 |
| NWD1 |  | EPB41L2 | ZNF502 |
|  |  | TCEAL3 | CORO6 |
|  |  | KLF15 | CNKSR2 |

### Integrative multi-omics analysis of UCEC

Three omics modalities were used for the MOFA factor analysis of UCEC, namely mRNA expression, miRNA expression, and DNA methylation. Six factor components were identified [**Figure 1**], representing independent underlying sources of variation. The process was robust to experiments with variation in algorithm initialization and data subsampling. In most of these experiments, Factors 1 and 2 were present, indicating their persistent significant roles in a variety of molecular layers.

**Figure 1:**
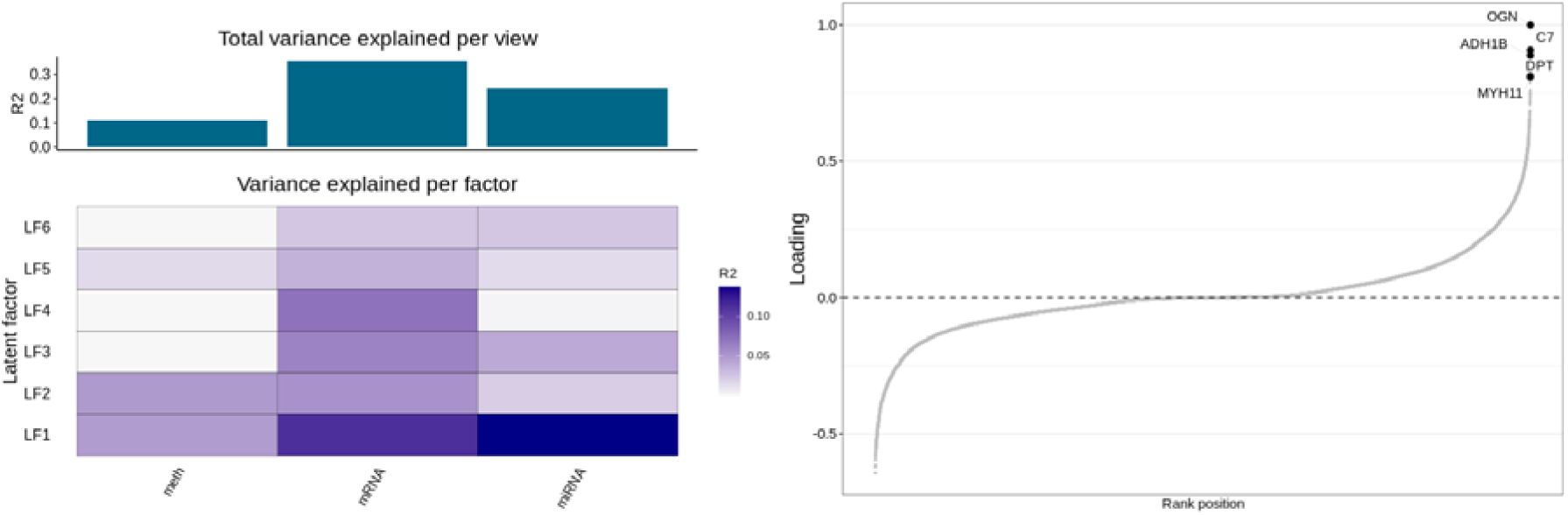
Proportion of total variance explained by individual factors, and cumulative proportion of total variance, in terms of R^2^, for each ‘view’ (i.e., modality) for UCEC; Plot of the mRNA features with largest loadings on Factor 1. The sign on the right indicates the direction of the loading.

Notably Factor 1 is active in the mRNA, miRNA, and methylation views. Analysis of the weights of strongly enriched genes for this factor in the context of the mRNA view revealed a set of genes involved in ECM organization, proteoglycans associated with ECM, muscle contraction, etc. [**Figure 2]**. It was interesting to note that Factor 5 was associated with cell cycle-related proteins.

**Figure 2:**
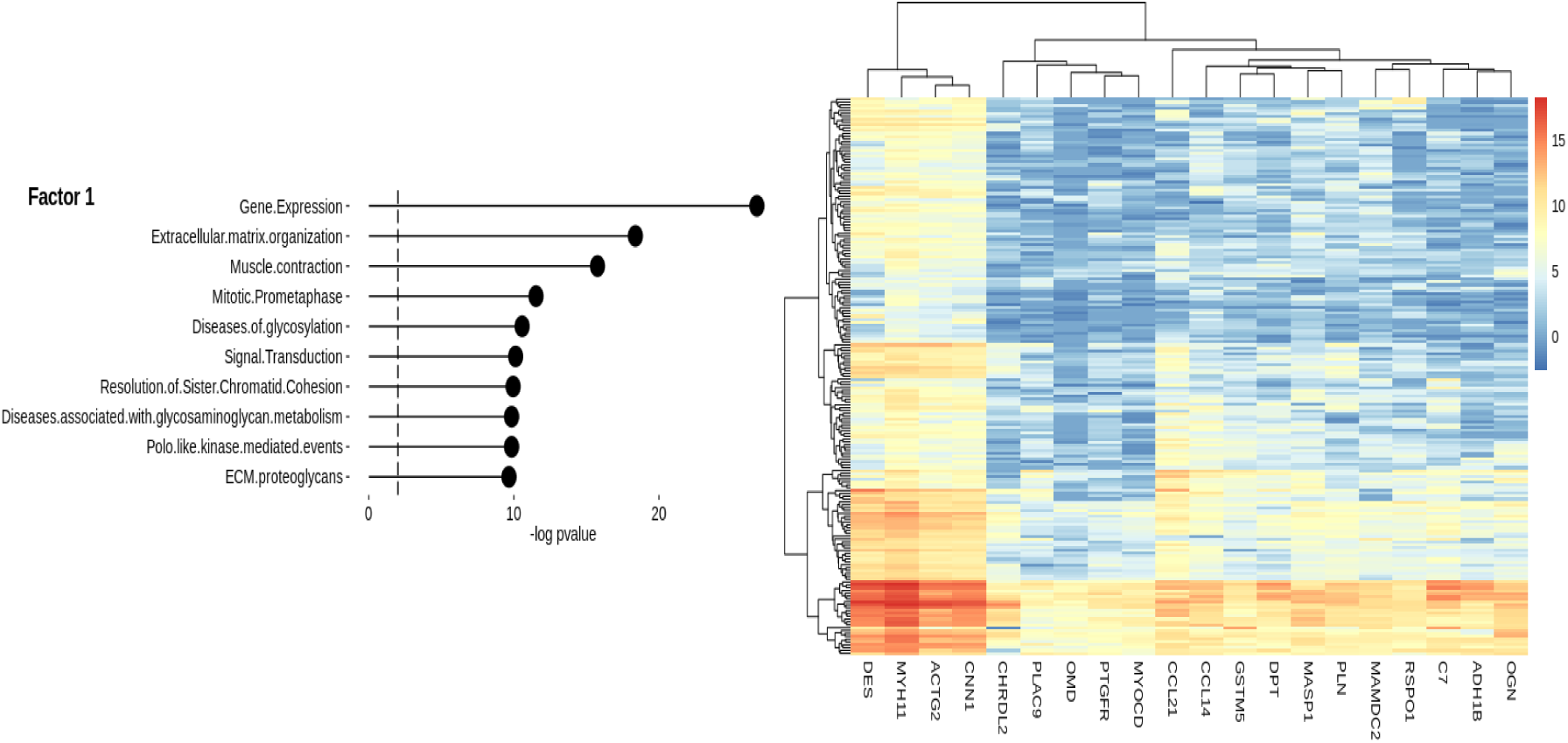
Characterization of Factor 1 of UCEC mRNA transcriptomics. (A) Gene set enrichment analysis (GSEA). (B) Heatmap of gene expression values for genes with the largest weights Given the persistent identification of Factors 1 and 2, we undertook a clustering analysis of the samples based on these Factors. A plot of the sample-wise loadings of these factors revealed a clustering of sample labels, specifically ‘Cancer’ vs ‘Control’ [**Figure 3**]. The clustering was nearly perfect, with four cancer samples misclassified in the cluster of ‘Controls’, and one control sample misclassified in the cluster of ‘Cancers’. These findings suggest the potential for using Factors 1 and 2 as the principal axes for a classification scheme, presently yielding an accuracy of 190/195 samples.

**Figure 3:**
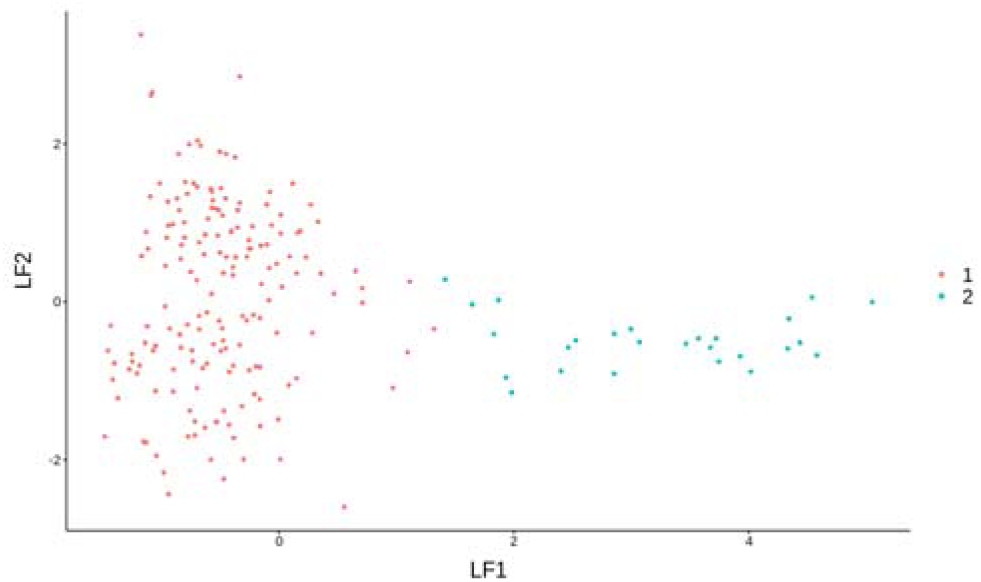
Visualization of samples using Factors 1 and 2 of UCEC mRNA modality. Red: cancer samples; blue: control samples.

Feature space reduction could be performed with MOVICS, and this was done using multiple techniques to determine a consensus feature space. Such techniques included those based on Median Absolute Deviation, Standard Deviation, Principal Components Analysis, and univariate Cox analysis. All three modalities namely mRNA, miRNA and methylation data were used to obtain technique-wise reduced feature spaces. A consistent consensus set of 53 up-regulated and 5 down-regulated genes was identified based on the results of the different feature selection methods in the different omics modalities.

Towards developing a multi-omic prognostic aid, it was sought to cluster the sample using consensus features from Cox univariate feature selection across the three modalities. Modality-wise, the Cox-analysis based feature selection yieleded 675 (/18540) mRNA features, 24 (/557) miRNA features, 14259 (/396064) cpg prob features. The number of subtypes was found by calculating the clustering prediction index (CPI) and gap-statistics. Four subtypes were unambiguously identified, namely CS1, CS2, CS3, and CS4, and found to be significantly associated with the overall survival (OS) rate (p-value ∼ 0.001; **Figure 4C**). Having identified the optimal *number* of clusters, an ensemble of ten clustering algorithms were applied to the dataset and consensus results were obtained, including consensus heatmap. The clustering results were checked with the specific patterns of expression across the mRNA and miRNA transcriptomes and with the patterns of epigenetic changes (**Figure 4**). Notably, cancer subtype 2 (CS2) is revealed as a high-risk group with significantly low survival, whereas the other three subtypes (CS1, CS3, and CS4) emerged as low-risk subtypes with significantly better survival. Some consensus features of the CS2 subtype include SLC39A5 and CLDN19. To assess the consistency of discovered subtypes with known classification schemes, we calculated the agreement between the existing information on cancer stage and the subtype class identified here (**Figure 5A**). From the alluvial diagram, it can be seen that CS3 subtype is enriched in early-stage UCEC (i.e., stages I and II), whereas CS2 is preferentially enriched in late-stage UCEC cancers (i.e., stages III and IV). This might also provide a basis for the survival differences between the two subtype cohorts. The genome-wide CNV changes were used to validate the subtypes, based on the fraction of genome alteration and the fraction of gain or loss in each sample presented in the identified subtypes (**Figure 5B)**. It is clear from these results that the differences between CS2 and CS3 subtypes extend significantly to the copy-number alteration profiles. It is seen that CS3 suffers significantly lower genomic gain or loss relative to that of CS2, explaining the profound differences in their overall survival. It remains to be further validated that the four subtypes identified here indeed match the clinical molecular subtypes, namely POLE ultramutated, microsatellite instability hypermutated, copy-number low, and copy-number high [10].

**Figure 4:**
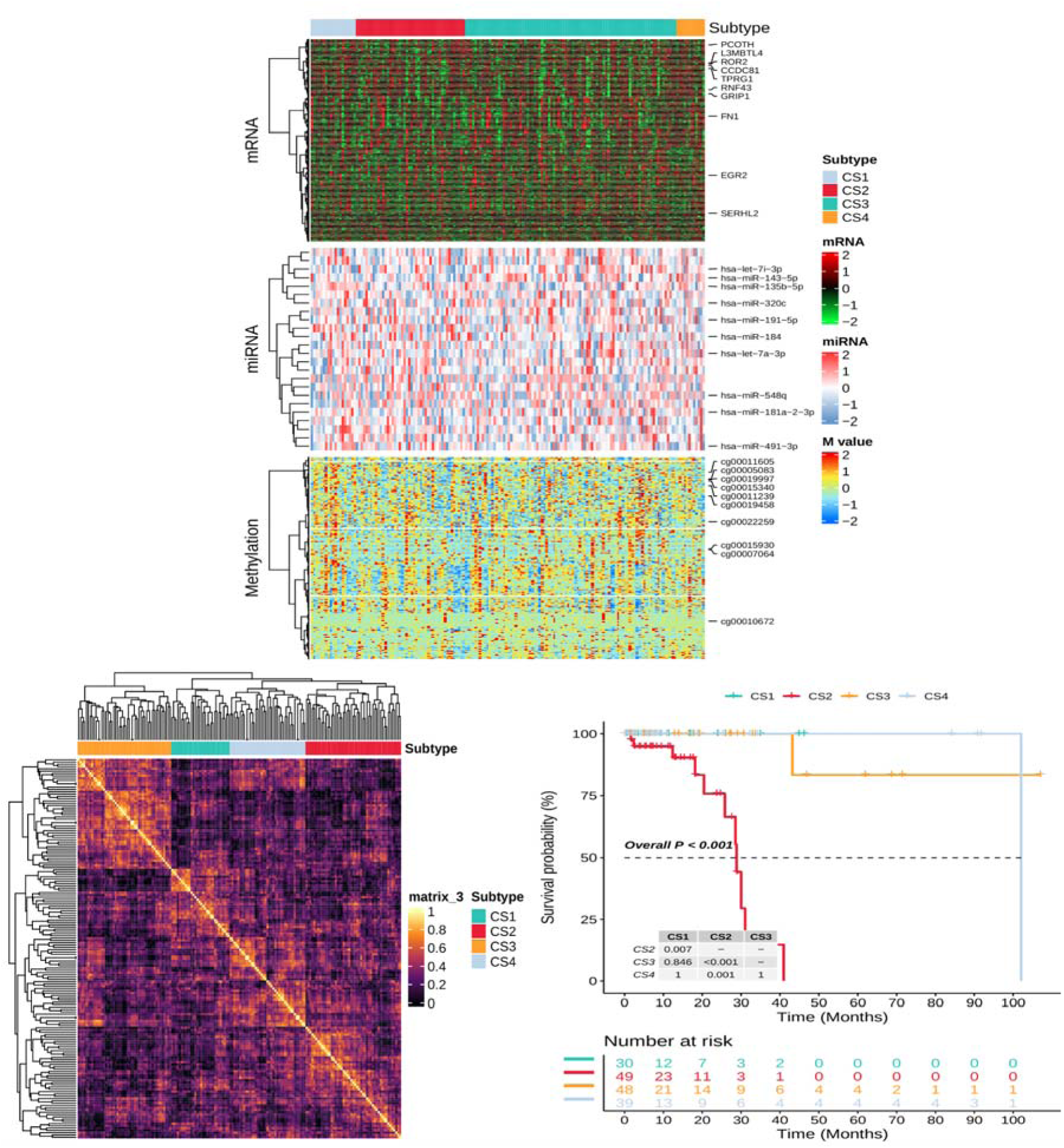
MOVICS multi-omics analysis of UCEC with Cox-based feature selection. (A) Integrative multi-omics analysis across samples using iClusterBayes; (B) Consensus heatmap based on results from 10 multi-omics integrative clustering algorithms with cluster number of 4; (C) Discovered consensus subtypes significantly associated with the overall survival Subtype-wise enrichment was quantified using the single-sample gene set enrichment analysis (ssGSEA) technique [**Figure 6**]. Interestingly, the CS2 subtype was enriched in positive regulation of phospholipid metabolic process, lipid kinase, membrane potential, cell-cell adhesion, etc. while CS3 subtype was enriched in cilium movement, and microtubule bundle formation.

**Figure 5:**
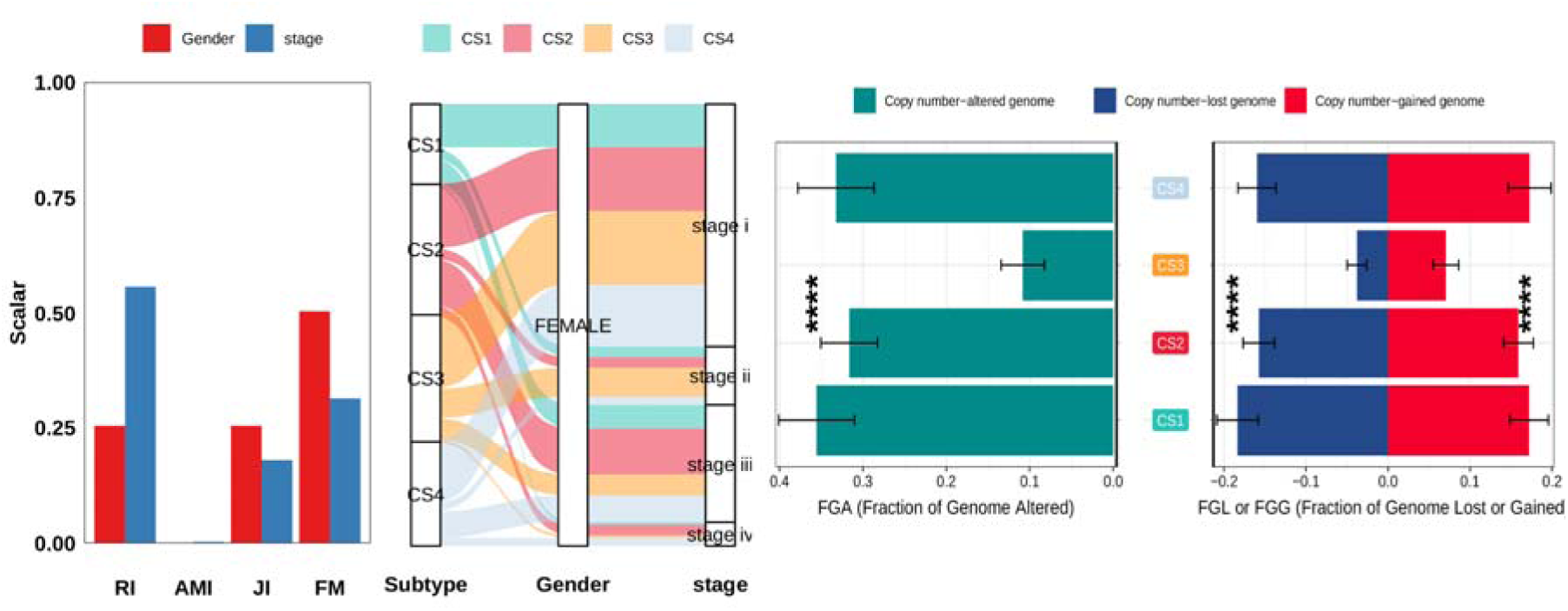
(A) Four measurements of agreement of the identified subtypes with AJCC staging system, namely Rand Index (RI), Adjusted Mutual Information (AMI), Jaccard Index (JI), and Fowlkes-Mallows (FM). (B) Barplot of fraction genome altered among 4 identified subtypes of UCEC.

**Figure 6:**
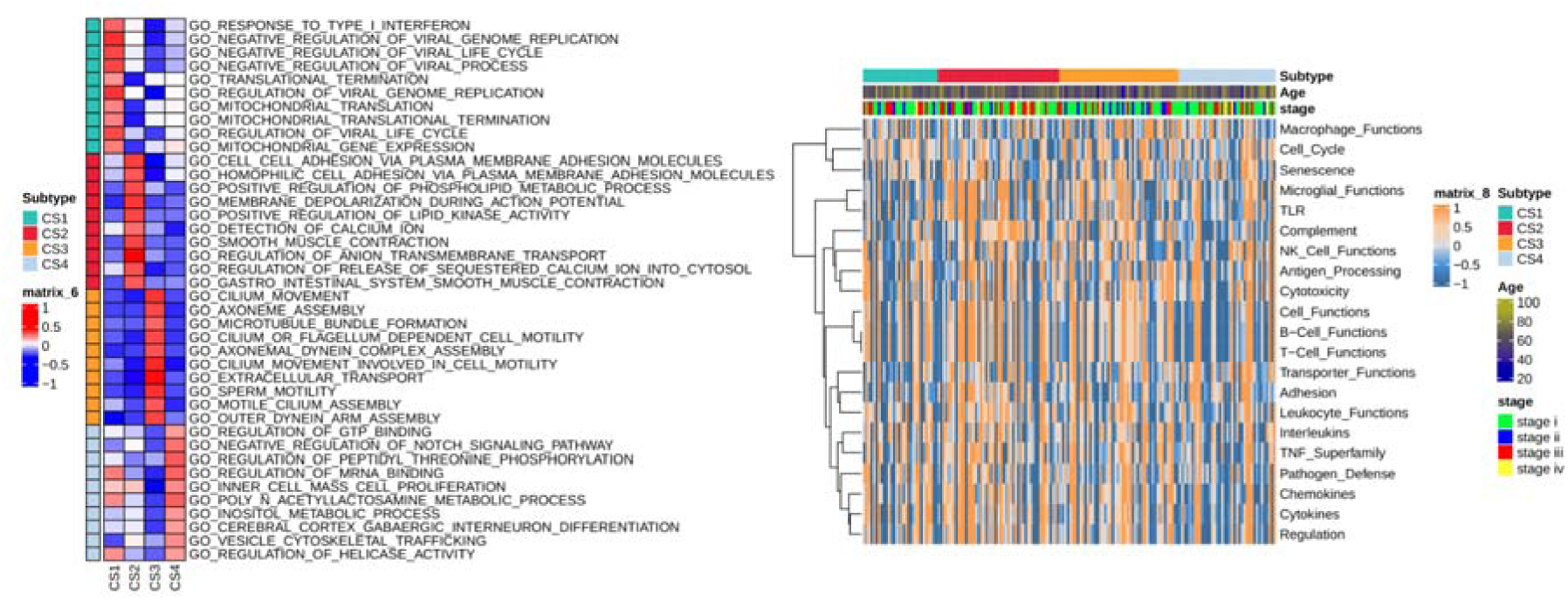
A. Single-sample geneset enrichment analysis for each subtype identified by MOVICS, highlighting the biological processes connected with the subtypes. CS2 and CS3 can be seen to show a distinct complement of enriched top pathways, which are oppositely regulated in the other subtypes. B. Heatmap of enrichment score of gene set of interest for the identified subtypes

### Integrative multi-omics analysis of OV

Three omics modalities were used for the MOFA factor analysis of ovarian cancer, namely mRNA expression, miRNA expression, and DNA methylation,. Six factor components were identified [**Figure 7**], representing independent underlying sources of variation. The process was robust to experiments with variation in algorithm initialization and data subsampling. In most of these experiments, Factors 1 and 2 recur, highlighting their general significance in multiple molecular layers. Factor 3 accounted exclusively for the methylation layer, indicating a distinct component of omics variation.

**Figure 7:**
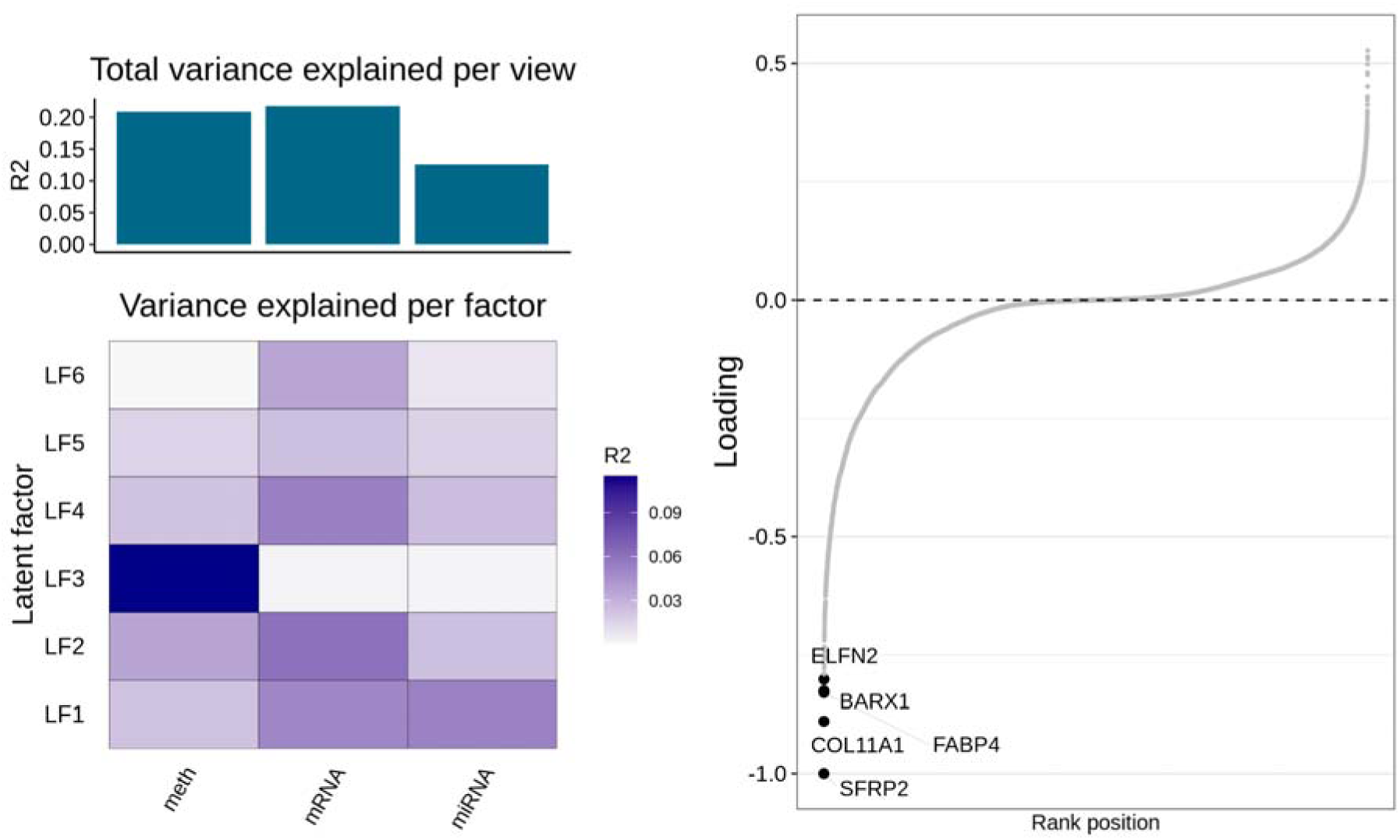
Left, Proportion of total variance explained by individual factors for each view (R^2^) and cumulative proportion of total variance explained for omics profiles in ovarian cancer. Right, Plot of the mRNA features with the largest loadings on Factor 1. The sign on the right indicates the direction of the loading.

Notably Factor 1 is active in the mRNA, miRNA, and methylation views. Analysis of th weights of strongly enriched genes for this factor in the context of the mRNA view revealed a set of genes involved in collagen biosynthesis and degradation as well as extracellular matrix associated processes [**Figure 8]**. It has been reported that collagen dysregulation plays a significant role in ovarian cancer progression [31].

**Figure 8:**
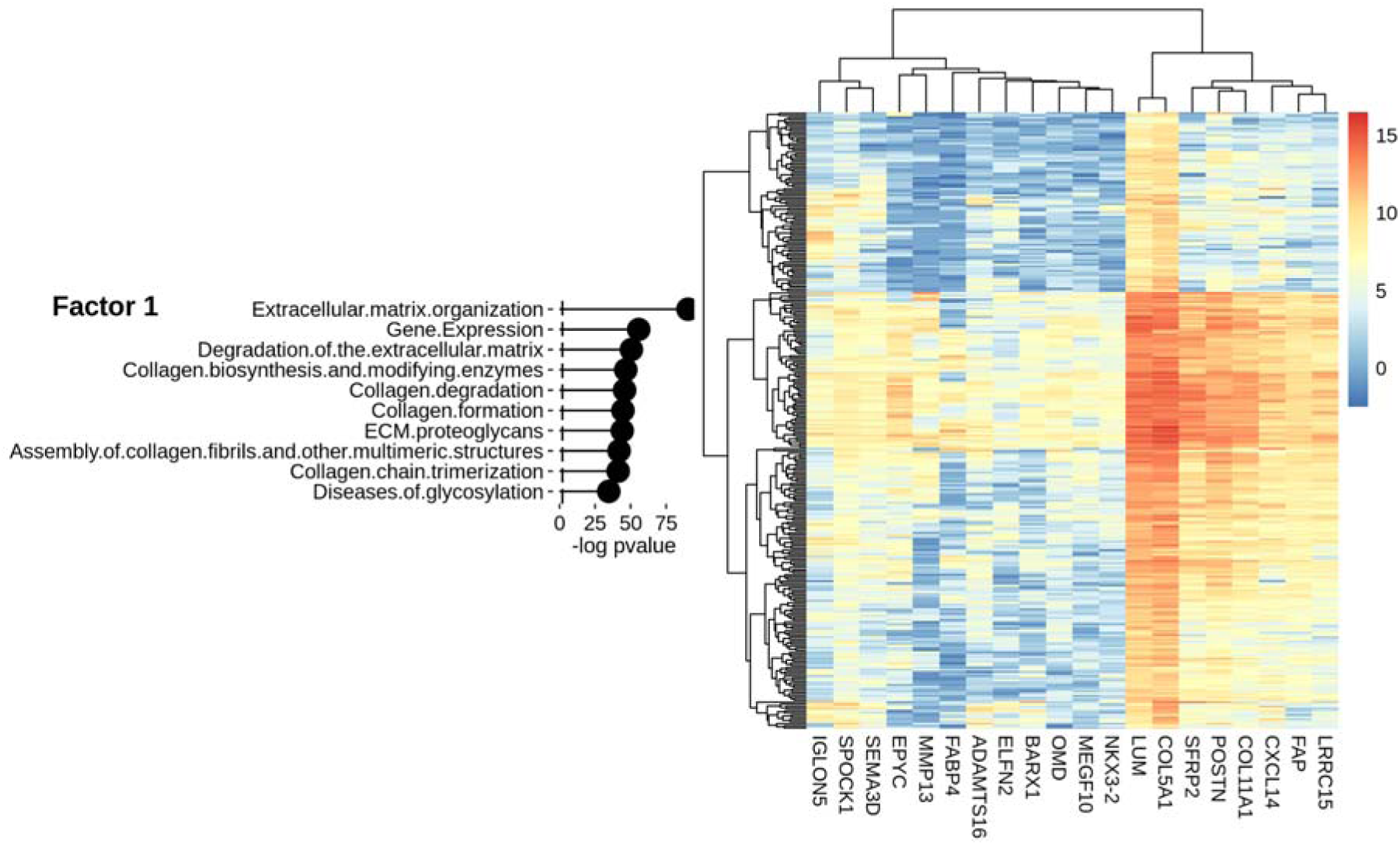
Characterization of Factor 1 in OV data for mRNA modality. (A) Gene set enrichment analysis (GSEA). (B) Heatmap of gene expression values for genes with the largest weights.

Feature space reduction could be performed with MOVICS, and this was done using multiple techniques to determine a consensus feature space. Such techniques included those based on Median Absolute Deviation, Standard Deviation, Principal Components Analysis, and univariate Cox analysis. All three modalities, namely mRNA, miRNA and methylation data were used to obtain technique-wise reduced feature spaces. A consistent consensus set of up-regulated and down-regulated genes was identified based on the results of the different feature selection methods in the different omics modalities.

Towards developing a multi-omic prognostic aid, it was sought to cluster the samples using consensus features from Cox univariate feature selection across the three modalities. Modality-wise, the Cox-analysis based feature selection yielded 675 (/18540) mRNA features, 24 (/557) miRNA features, 14259 (/396064) cpg probe features. The number of subtypes was found by calculating the CPI and gap-statistics and evaluating the evidence from literature. Three subtypes were unambiguously identified, namely CS1, CS2, and CS3, possibly corresponding with germ cell tumors (teratomas and dysgerminomas), stromal tumors (granulosa tumors) and sarcomas. The prognostic subtype identification was found to be significantly associated with the OS rate (p-value ∼ 0.001; **Figure 9C**). Having identified the optimal *number* of clusters, an ensemble of ten clustering algorithms were applied to the dataset and consensus results were obtained, including consensus heatmap. The clustering results were checked with the specific patterns of expression across the mRNA and miRNA transcriptomes and with the patterns of epigenetic changes (**Figure 9**). Notably, cancer subtype 1 (CS1) showed the best survival rates, whereas cancer subtype 2 (CS2) and cancer subtype 3 (CS3) tended to show a similar survival curves with significantly higher risk and lower survival. To assess the consistency of discovered subtypes with known classification schemes, we calculated the agreement between the existing information on cancer stage and the subtype class identified here (**Figure 10A)**. From the alluvial diagram, it can be seen that stage-II OV cancer is enriched in CS1 subtype, whereas CS2 is preferentially enriched in stage-III OV cancers. This might also provide a basis for the survival differences between the two subtype cohorts. The genome-wide CNV changes were used to validate the subtypes, based on the fraction of genome alteration and the fraction of gain or loss in each sample presented in the identified subtypes (**Figure 10B)**. It is clear that the differences between CS1 and CS2 subtypes extends significantly to the copy-number alteration profiles. In particular, it is seen that CS2 suffers significantly higher genomic gain and loss (with significantly higher fraction of altered genome) relative to that of CS2, explaining the profound differences in their overall survival. It remains to be further validated that the three subtypes identified here might correspond with the indicated clinical molecular subtypes.

**Figure 9:**
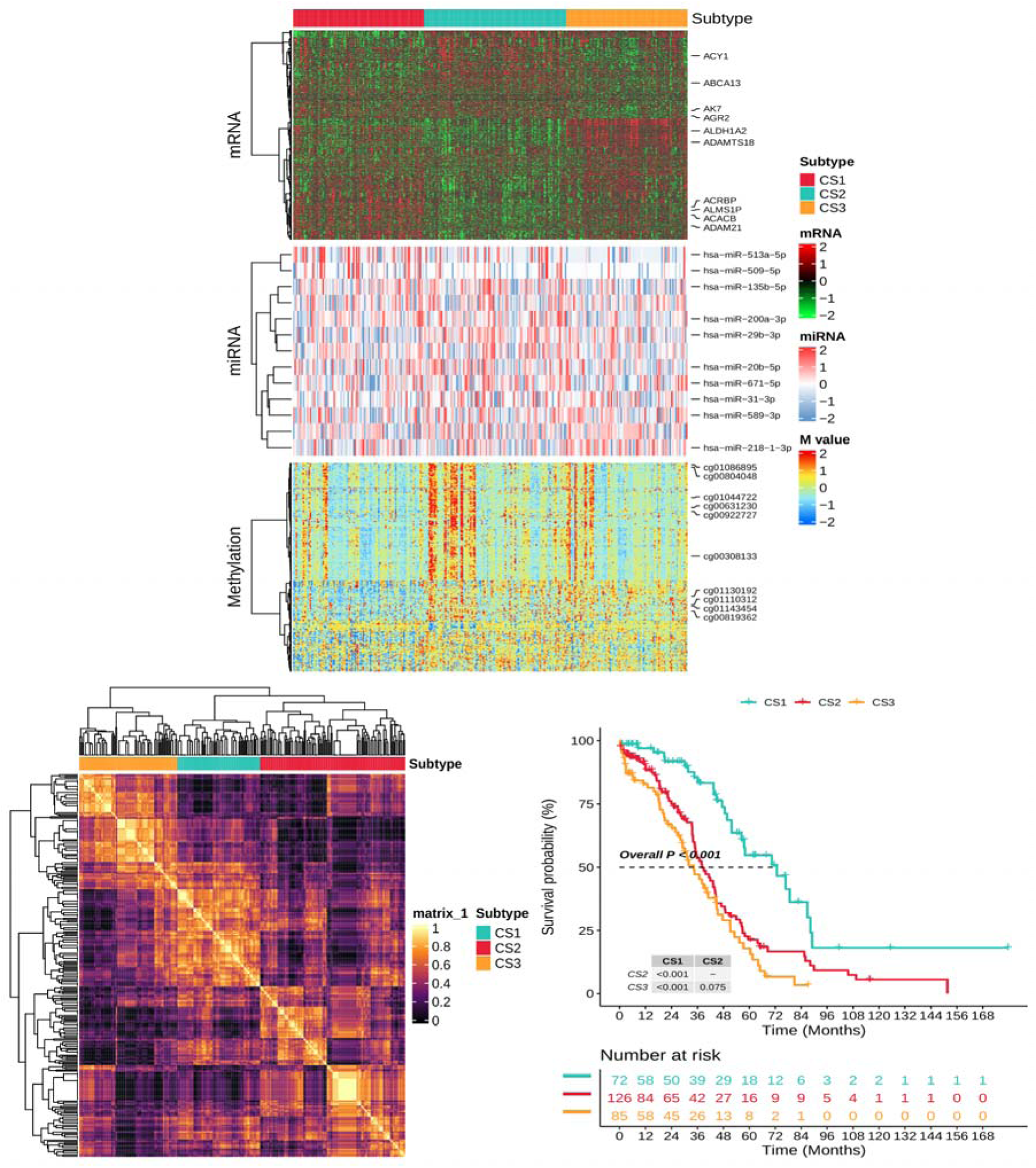
MOVICS multi-omics analysis of OV with Cox-based feature selection. (A) Integrative multi-omics analysis across samples using iClusterBayes; (B) Consensus heatmap based on results from 10 multi-omics integrative clustering algorithms with cluster number of 3; (C) Discovered consensus subtypes significantly associated with the overall survival Subtype-wise enrichment was quantified using the single-sample gene set enrichment analysis (ssGSEA) technique [**Figure 11**]. Interestingly, the CS3 subtype was enriched in collagen fibril organization and metabolic process, sprouting angiogenesis, endothelial cell migration, etc.

**Figure 10:**
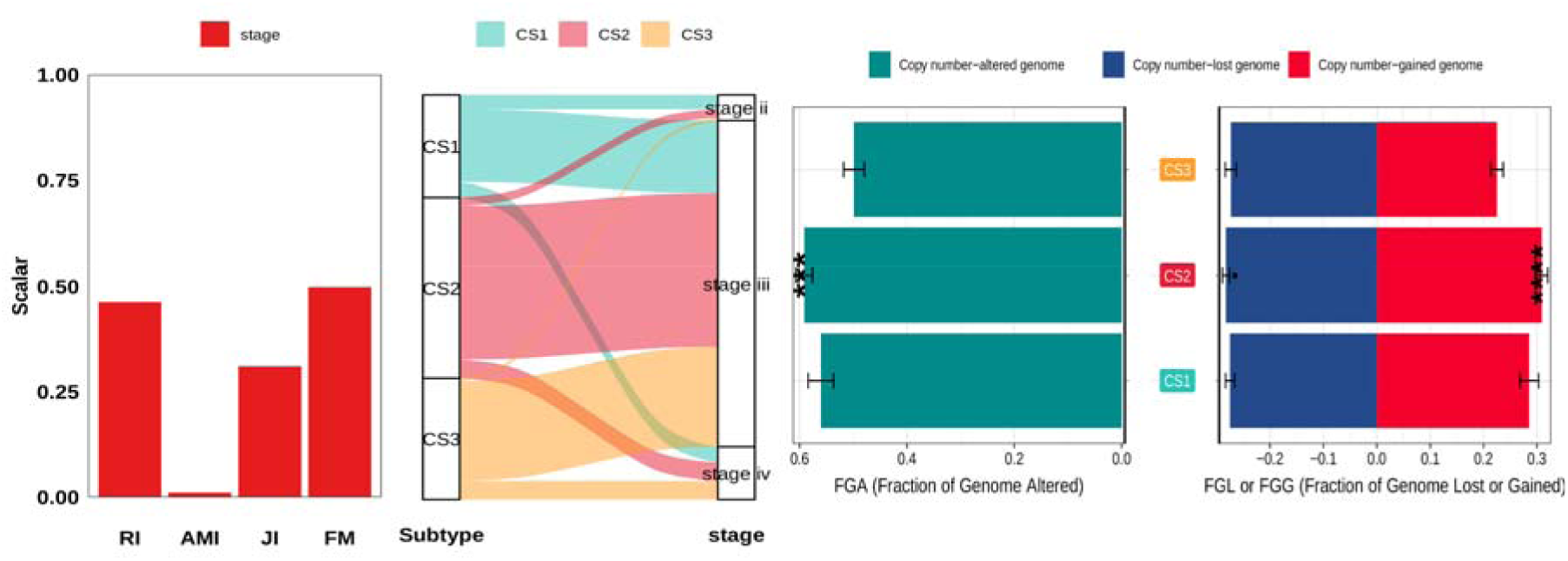
(A) Four agreement measurements, including Rand Index (RI), Adjusted Mutual Information (AMI), Jaccard Index (JI), and Fowlkes-Mallows (FM).of identified subtypes with the AJCC staging system. (B) Barplot of fraction genome altered among three identified OV subtypes.

**Figure 11:**
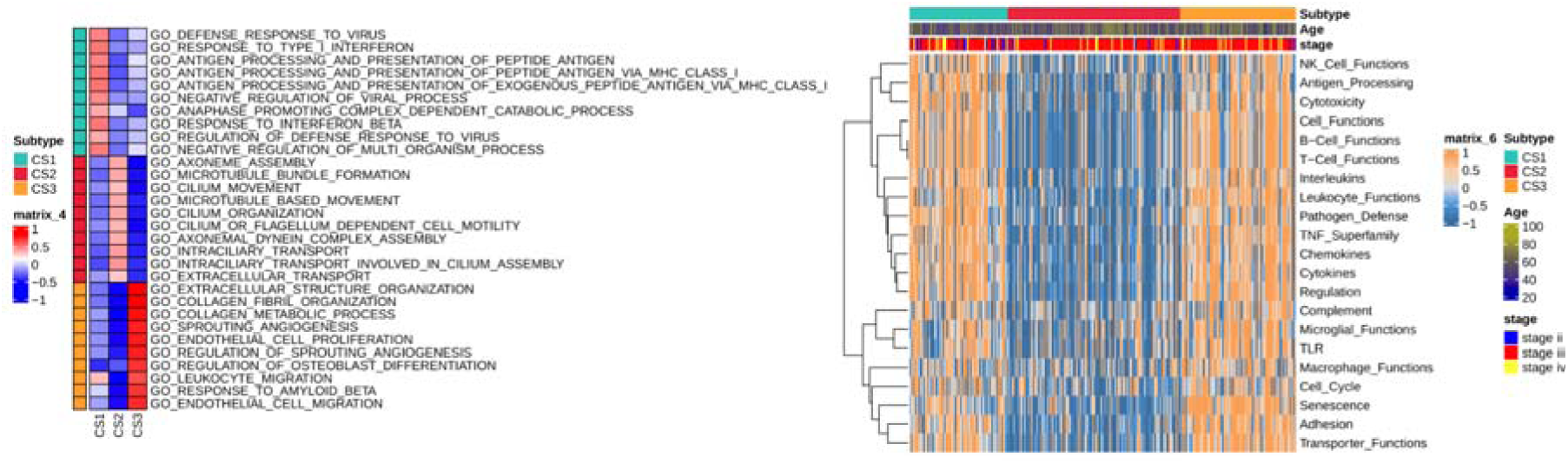
A. Single-sample geneset enrichment analysis for each subtype identified by MOVICS, highlighting the biological processes connected with the subtypes. CS2 and CS3 can be seen to show a distinct complement of enriched top pathways, which are oppositely regulated in the other subtypes. B. Heatmap of enrichment score of gene set of interest for the identified subtypes

### Integrative bi-omics analysis of CESC

Figure 12A depicts the correlation circle plots for the rCCA-based shrinkage method using a cutoff of 0.5. Canonical correlation analysis showed the main correlation between mRNA and miRNA in a two-component plot, for example, MEG3 with hsa-miR-145-3p and other miRNAs. The relationship between mRNA and miRNA profiles was also visualized with a relevance network based on an absolute correlation threshold > 0.5 (Figure 12B). A complex correlation structure is seen, in which three clusters could be identified: a singleton gene, a group of six genes, and a group of twelve genes. Hsa-miR-145-3p appeared to act as a bridge between the groups of six and twelve genes. The network exhibited edges with large positive correlations between the nodes.

**Figure 12:**
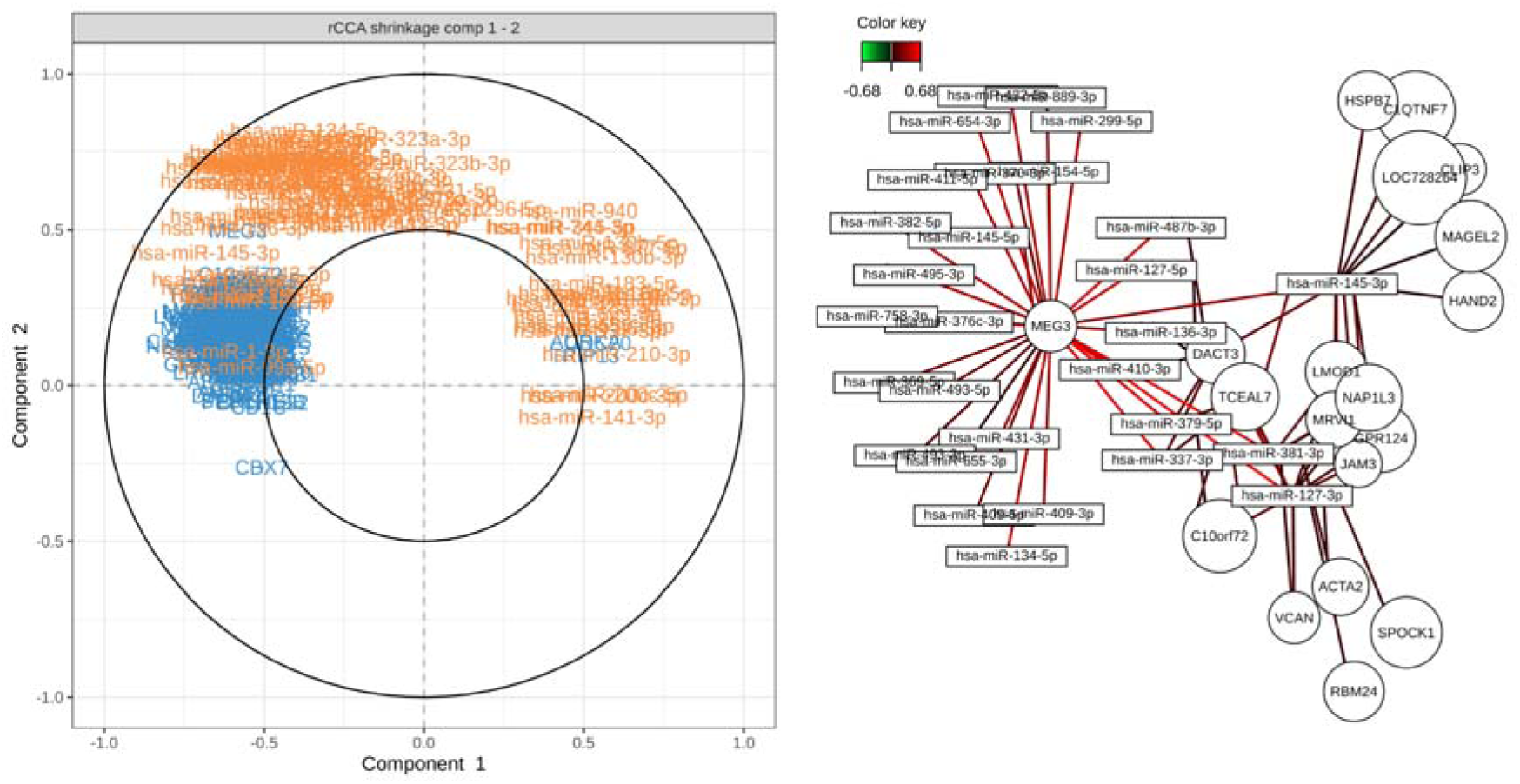
(A) Correlation circle plots derived from rCCA applied to CESC data, illustrating the relationship between miRNA and mRNA expression. (B) The relevance network plot of the mRNA nodes showing every bipartite relationship (mRNA-miRNA) with an absolute correlation value > 0.5. Genes are represented by the circular nodes, and miRNA by rectangles. The color of the connecting lines (edges) indicates the strength of the bipartite links.

A correlation circle plot derived from the sPLS analysis identifies the factors explaining the overall covariance between the two datasets (**Figure 13A). Figure 13B** shows the relevance network based on a threshold absolute correlation > 0.7. Two subnetworks could be discerned: one, comprising a central hsa-miR-205-3p that is mostly negatively linked with thirteen genes; and two, comprising two miRNA features (hsa-miR-944 and hsa-miR-205-5p) and ∼ 25 genes in a bipartite structure with largely positive weighted links.

**Figure 5.13:**
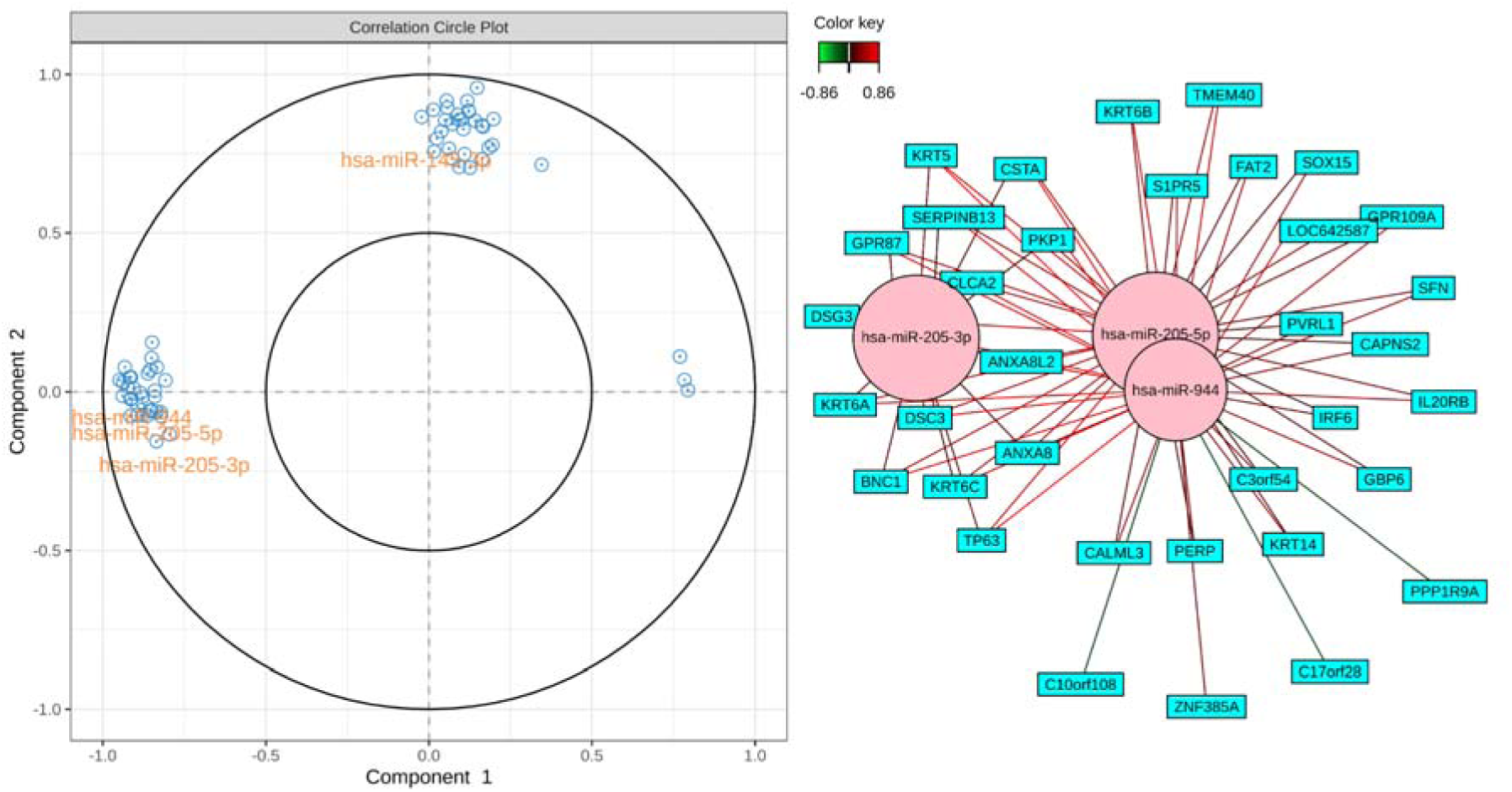
(A) Correlation circle plot from the sPLS performed on the CESC data. (B) The bipartite relevance networks connecting a gene (rectangle) to an miRNA (circle) if absolute correlation in similarity matrix > 0.7.

To obtain further clarity on the feature structure from the sPLS analysis, a Cluster Image Map (CIM) of mRNA vs miRNA expression could be visualized. An absolute correlation threshold > 0.7 was used to obtain significant miRNA and mRNA features.

From Figure 14, two clusters of miRNA expression could be seen: each cluster negatively regulating an exclusive set of mRNAs. This structure is also reflected in the corresponding mRNA clustering pattern. Given these striking results, it may be helpful to observe the composition of the miRNA clusters; cluster 1: hsa-miR-205-5p, hsa-miR-205-3p, hsa-miR-944, and hsa-miR-203a-3p; and cluster 2: hsa-miR-133a-3p, hsa-miR-1-3p, hsa-miR-145-5p, and hsa-miR-145-3p.

**Figure 14:**
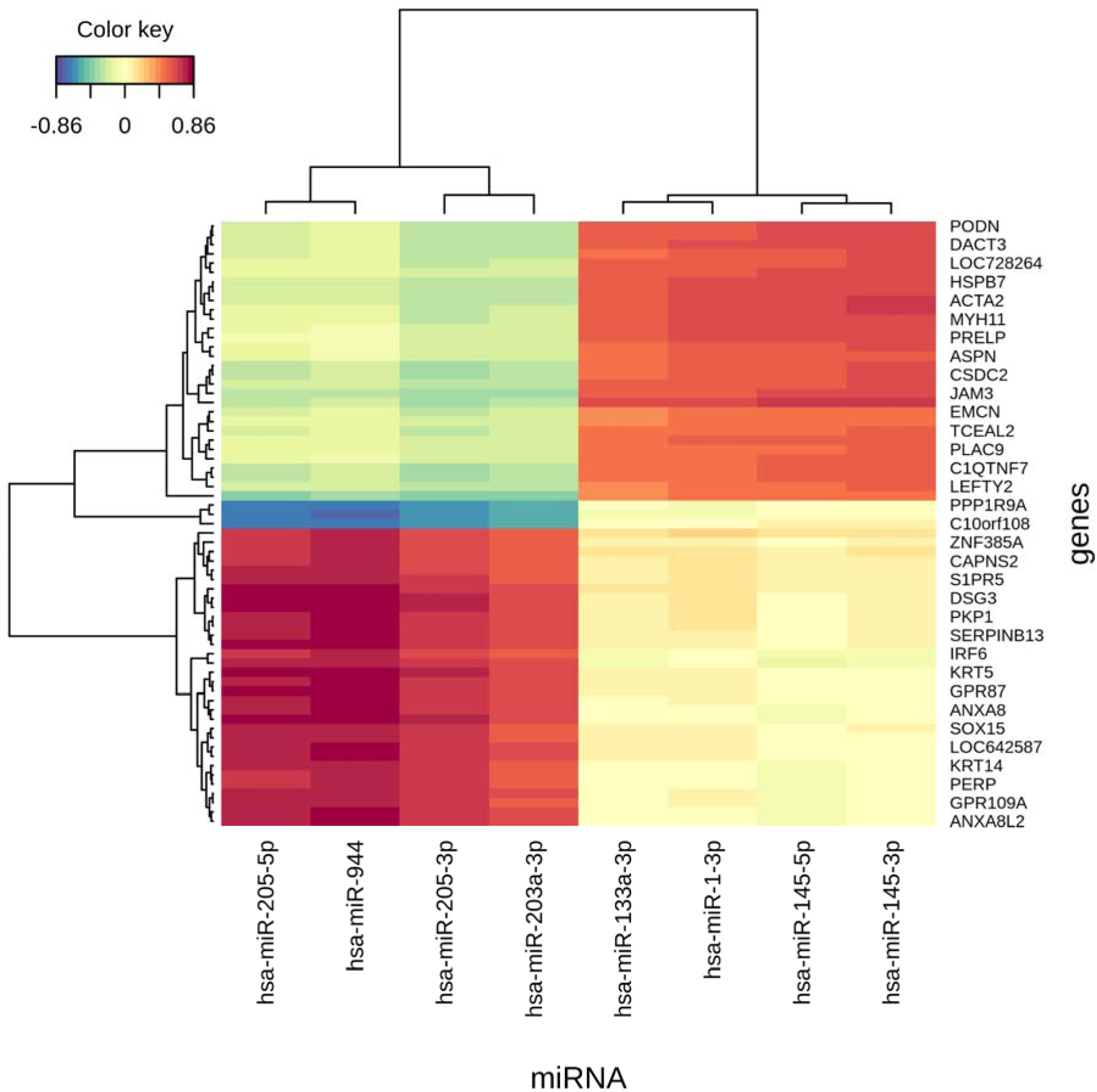
Clustered Image Map generated from sPLS analysis of CESC mRNA and miRNA transcriptomics based on an absolute correlation threshold > 0.7. Distance between chosen features was calculated using the Euclidean measure.

### Consensus of stage-salient and multi-omics biomarkers

In the foregoing, we have identified stage-salient biomarkers for each cancer and followed it up with a multi-omics analysis. The multi-omics analysis of each cancer has in turn identified multiple potential biomarkers. In the below, we sought to determine the consensus between the two sets of biomarkers for each cancer toward furnishing a more robust set of potential biomarkers for each of the gynecological cancers. With respect to UCEC, the consensus of MOVICS analysis and stage-salient linear model yielded the following eight consensus biomarkers: one stage-2 salient (KLK12), four stage-3 salient (SLC39A5, CLDN19, PNOC, RSPO4), and three stage-4 salient (MAGEA9B, KIF1A, CBS) genes. The stage-wise expression distribution of these eight genes is depicted in Figure 15. The consensus of MOFA factor analysis and stage-salient linear model yielded the following 19 consensus biomarkers: three stage-1 salient (CYP1B1, PPP1R3C, LRCH2), and 16 stage-4 salient (DNASE1L3, CPA3, MYOZ2, RBM20, COL14A1, ADAMTSL1, ANO4, FCER1A, FGF14, LOC284276, AFF3, PDZRN3, FAM78B, C6orf138, ITGA9, CD1C) genes. The stage-wise expression distribution of these 19 genes is depicted in Figure 16.

**Figure 15:**
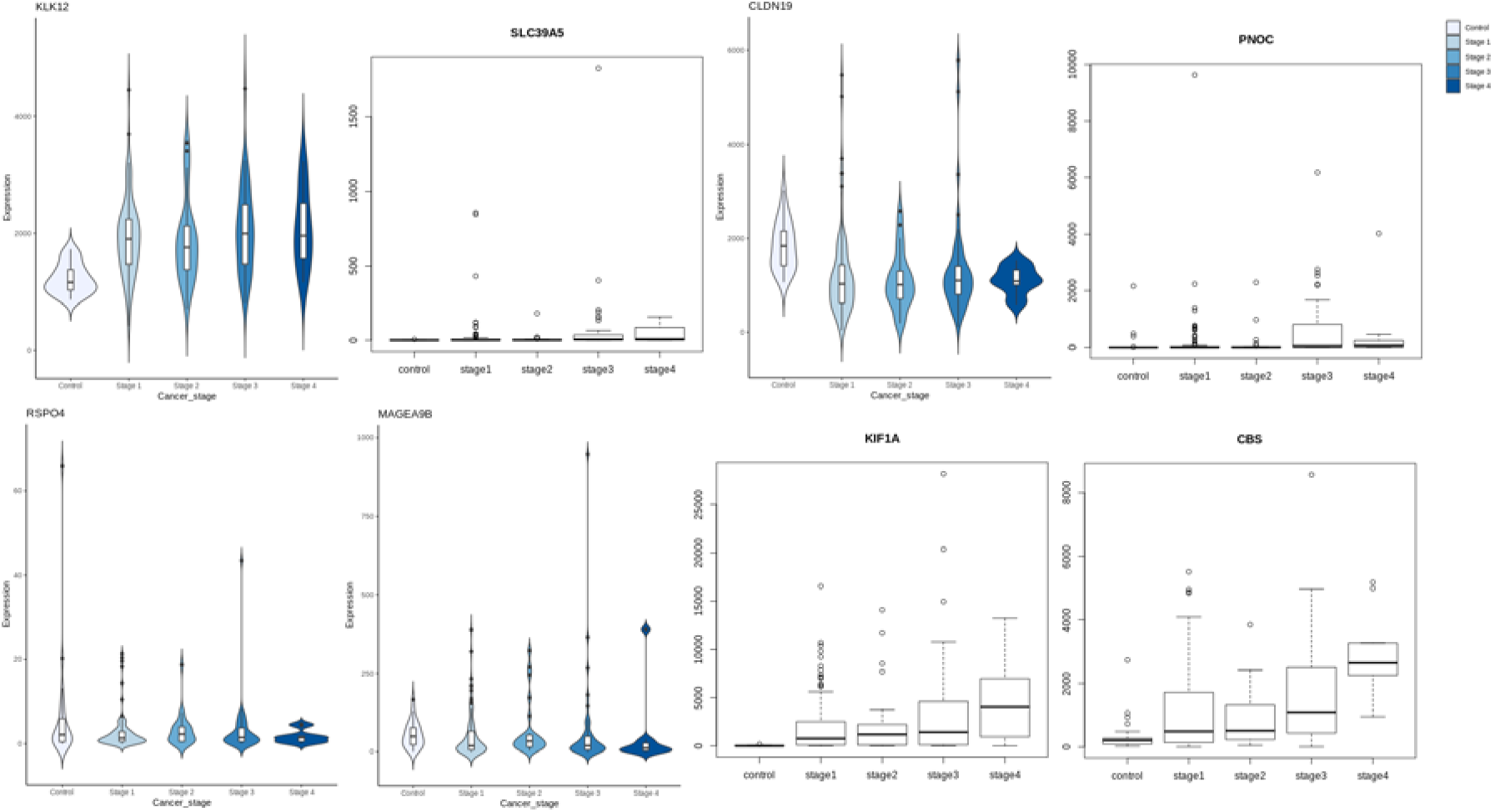
Stage-wise distribution of expression of UCEC feature genes at the consensus of MOVICS multi-omics analysis and stagewise linear models. It is seen that KLK12, SLC39A5, PNOC, RSPO4, MAGEA9B, KIF1A, and CBS were overexpressed, whereas CLDN19 was the sole downregulated feature.

**Figure 16:**
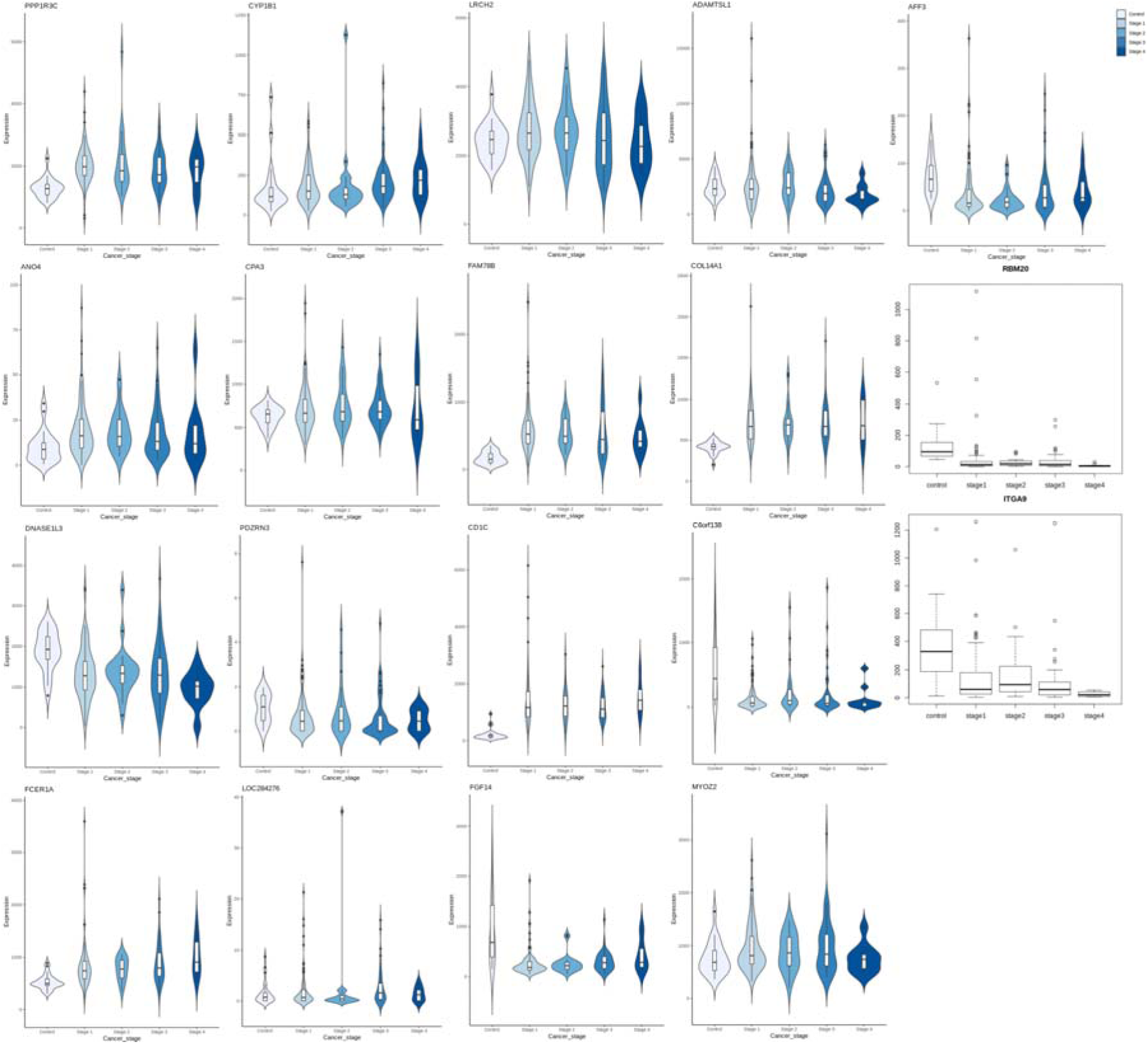
Stage-wise distribution of expression of UCEC feature genes at the consensus of MOFA factor analysis and stagewise linear models. Genes were both upregulated, like LRCH2, and downregulated, like PDRZN3.

With respect to OV, the consensus of MOVICS analysis and stage-salient linear model yielded just one consensus biomarker: C7 (stage-1 salient). The consensus of MOFA factor analysis and stage-salient linear model yielded the following nine consensus biomarkers: AGPAT3, AUTS2, CETN3, GNG12, LONRF2, LYSMD3, MBLAC2, NOTCH2, SH2D4A (all stage-1 salient). The stage-wise expression distribution of these ten genes is depicted in Figure 17.

**Figure 17:**
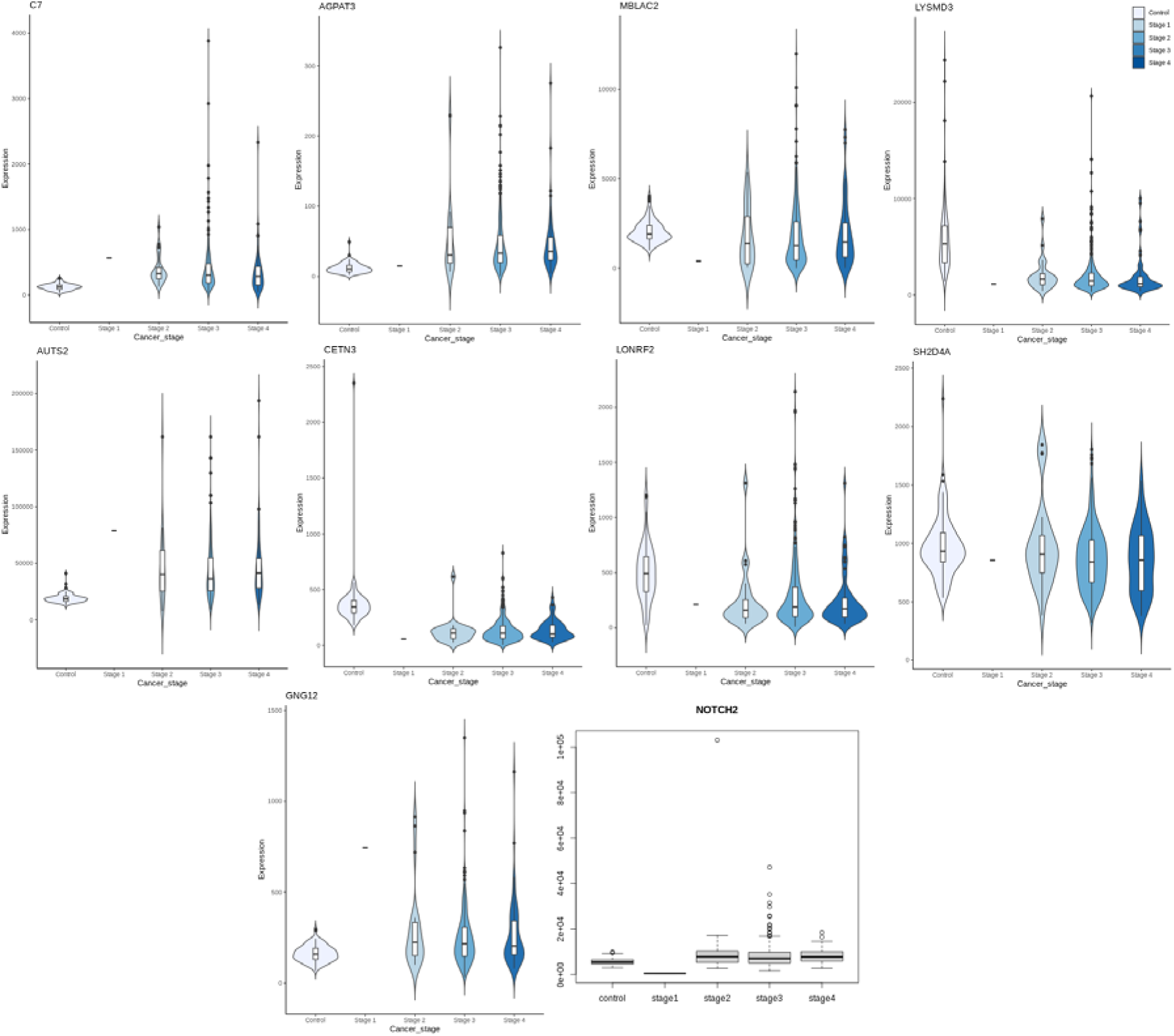
Stage-wise distribution of expression of OV feature genes at the consensus of MOFA factor analysis and stagewise linear models. Genes were both upregulated, like GNG2, and downregulated, like CETN3.

With respect to CESC, the consensus of bi-omics analysis and stage-salient linear model yielded just one consensus biomarker: HAND2 (stage-4 salient). The consensus of canonical correlation analysis and sPLS analysis yielded the following five biomarkers: DACT3, HSPB7, ACTA2, JAM3, C1QTNF7. The stage-wise expression distribution of these six genes is depicted in Figure 18.

**Figure 18:**
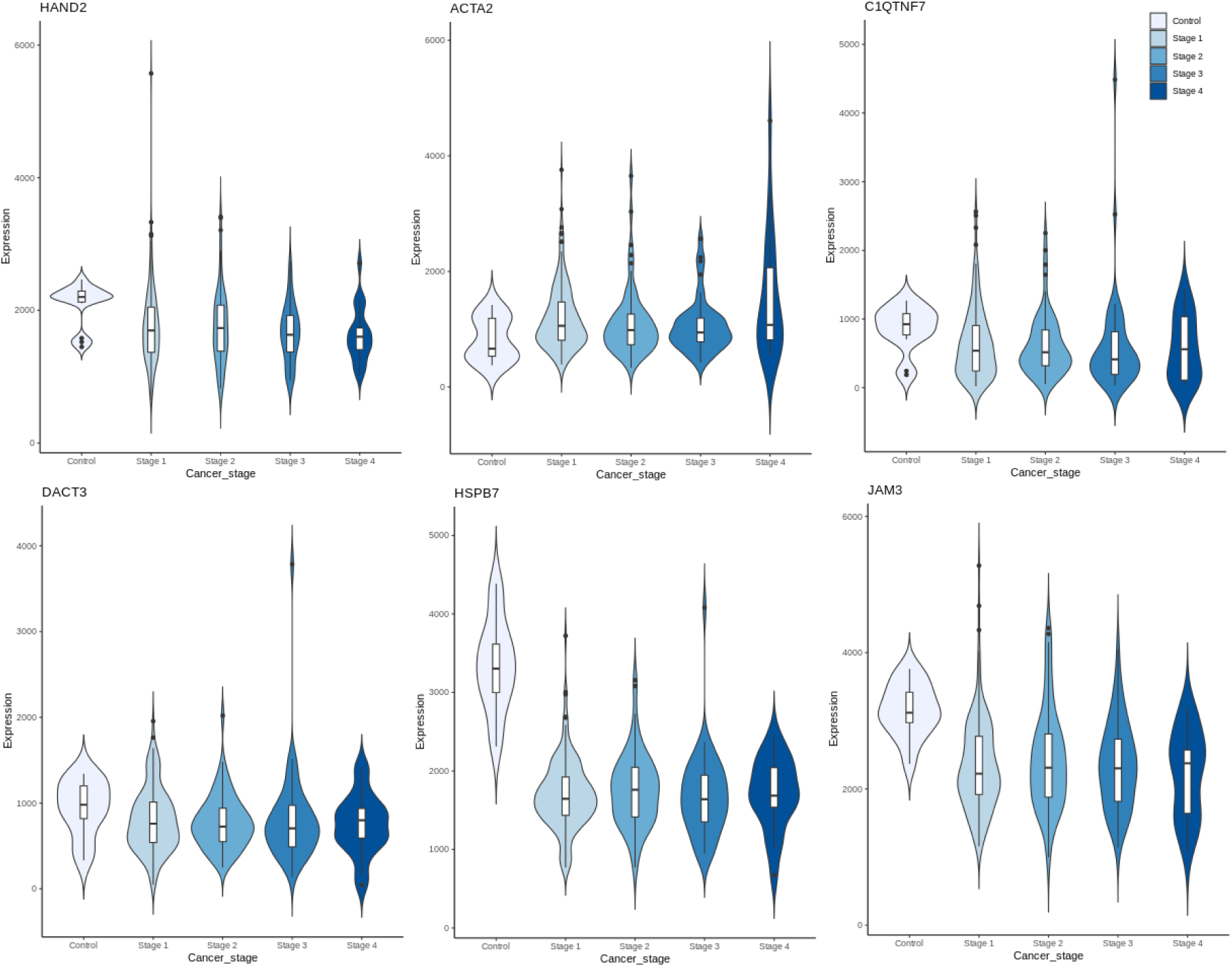
Stage-wise distribution of expression of CESC feature genes at the consensus of biomics analysis and stagewise linear models. Genes were both upregulated, like ACTA2, and downregulated, like HAND2 and HSPB7.

## DISCUSSION

Given the extended burden of disease in gynecological cancers, it is imperative to improve access to care in resource-limited settings. There has been notable progress in the treatment of disease reported in recent literature. AI-assisted screening methods have demonstrated promising results in improving cervical cancer detection rates while reducing workload [32]. Novel immunotherapy combinations, particularly with pembrolizumab, have shown improved survival in advanced cervical cancer [33]. Liquid biopsy techniques for early detection of ovarian cancer show promising results in detecting circulating tumor DNA [34]. PARP inhibitors have emerged as a cornerstone of ovarian cancer treatment, especially in BRCA-mutated cases [35]. Novel combinations of immunotherapy with anti-angiogenic agents have demonstrated improved progression-free survival in platinum-resistant ovarian cancer [36]. With respect to endometrial cancers, immunotherapy has shown remarkable success in MMRd tumors, with response rates exceeding 50% in some studies [37,53]. Novel combinations of targeted therapies based on molecular profiling have demonstrated improved outcomes in advanced endometrial cancer [38].

In this context, our findings could inform the design of early-stage diagnostic models and meaningfully advance tailored treatment programs [39]. Here, we provide a discussion of the consensus candidate biomarkers identified for each of the gynecological cancers in the light of the available literature. This provides support for the identified biomarkers as well as enables us to ascertain their novelty. Further an ROC curve analysis is performed for each individual candidate biomarker for their ability to discriminate between respective cancer and normal samples. Biomarkers with AUROC > 0.9 are called singular biomarkers.

### Literature review of consensus biomarkers

KLK12 has been reported for modulating proangiogenic effect [40], the bioavailability of growth factors [41], and regulating cell adhesion and migration [42]. There have been reports of its overexpression in UCEC patients [43], which is reaffirmed by the results here. RSPO4 is a circadian rhythm-related factor that might flag the onset and development of endometrial cancer [44] and observed to be upregulated here.

Overexpression of MAGEA9B with significant gain of copy number was found to be associated with worse overall survival [45], which accords with the upregulation observed here. CYP1B1 has been linked to notable alterations in estrogen metabolism that underlie inter-individual variations in the risk of endometrial cancer [46].

C7 has been reported as upregulated in ovarian cancers [47,48]. Similar findings were observed here. Hypermethylated HAND2 is known to be essential for the development, morphogenesis, and control of angiogenesis, and has been used in a nine-gene panel for the screening and early diagnosis of cervical cancer [49]. JAM3 had been earlier identified as a methylation marker of cervical cancer specific to maintenance of polarity in epithelial and endothelial cells [50] and is observed downregulated here.

A substantial increase in HSPB7 promoter methylation has been observed in nine cancer types including cervical cancer. Of these, three cancer types—cervical cancer, sarcoma, and glioblastoma—showed significant copy number gains in HSPB7, with gains of 0.15, 0.14, and 0.07 copies, respectively [51]. C1QTNF7 is a known differentially expressed protein associated with cervix small cell cancer [52], observed downregulated here.

### Singular biomarkers from ROC curve analysis

ROC curve analysis was used to determine the diagnostic value of consensus biomarker features identified for each of the gynecological cancers. This included a total of 27 biomarker features for UCEC, 10 biomarker features for OV, and 6 biomarker features for CESC. Each feature was separately assessed for the area under the ROC curve (AUC) as well as sensitivity and specificity. Biomarker features with AUROC >0.9 were deemed capable of discrimination between cancer and normal and could be further explored in biomarker panels for early-stage cancer screening. Such biomarker features, termed singular biomarkers, are displayed in **Table 4**. and their ROC curves for the respective cancers shown in Figure 19, Figure 20, and Figure 21. For UCEC, it is seen that eleven features exceed 0.9 AUROC, namely MYOZ2, CYP1B1, PPP1R3C, DNASE1L3, ADAMTSL1, LRCH2, RBM20, LOC284276, FAM78B, COL14A1, and PDZRN3. For OV, it is seen that just two features exceed 0.9 AUROC, namely C7 and LONRF2. For CESC, it is seen that all six features exceed 0.9 AUROC, with HAND2, C1QTNF7 and JAM3 exceeding 0.99 AUROC.

**Figure 19:**
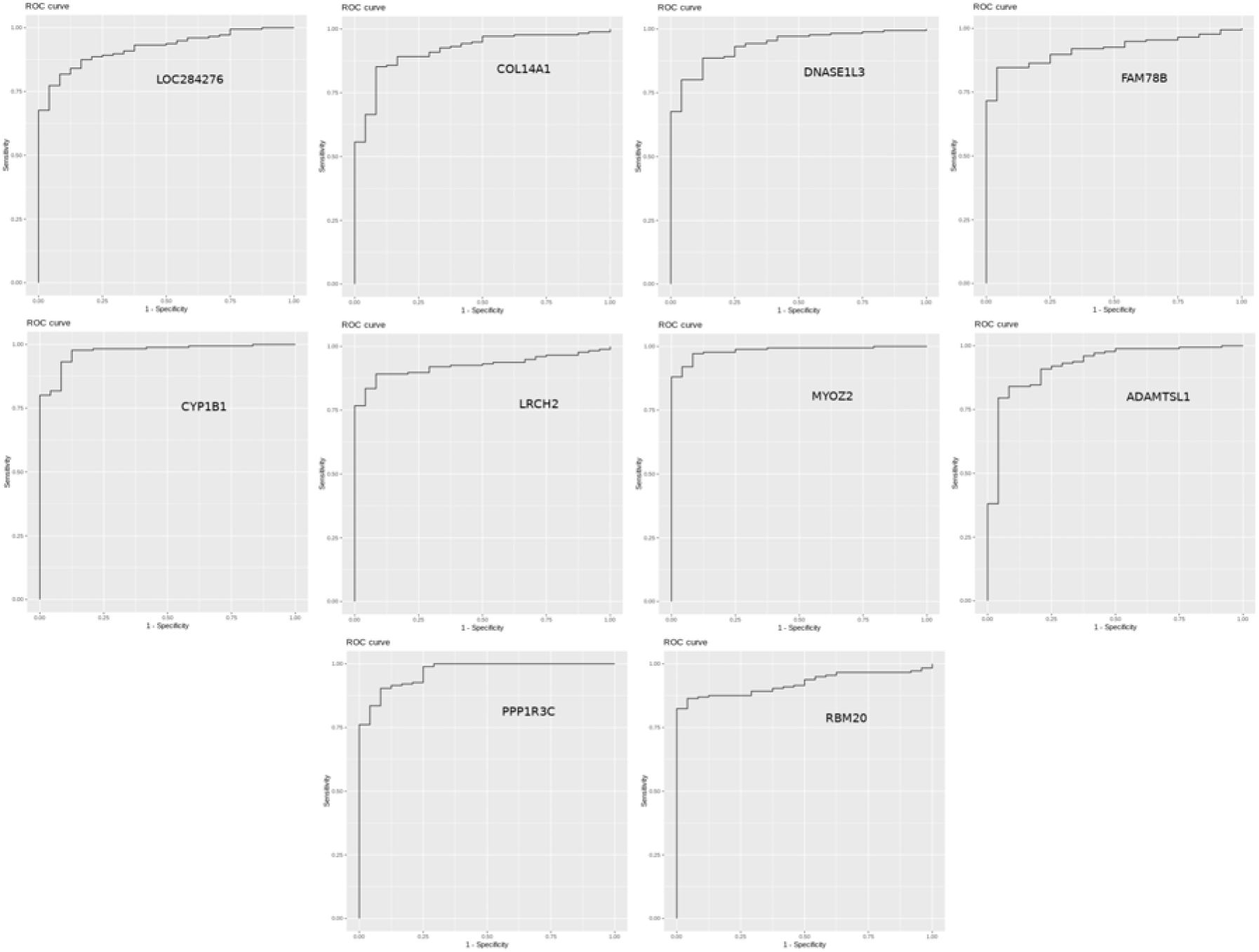
ROC curves for the top 10 UCEC singular biomarkers.

**Figure 20:**
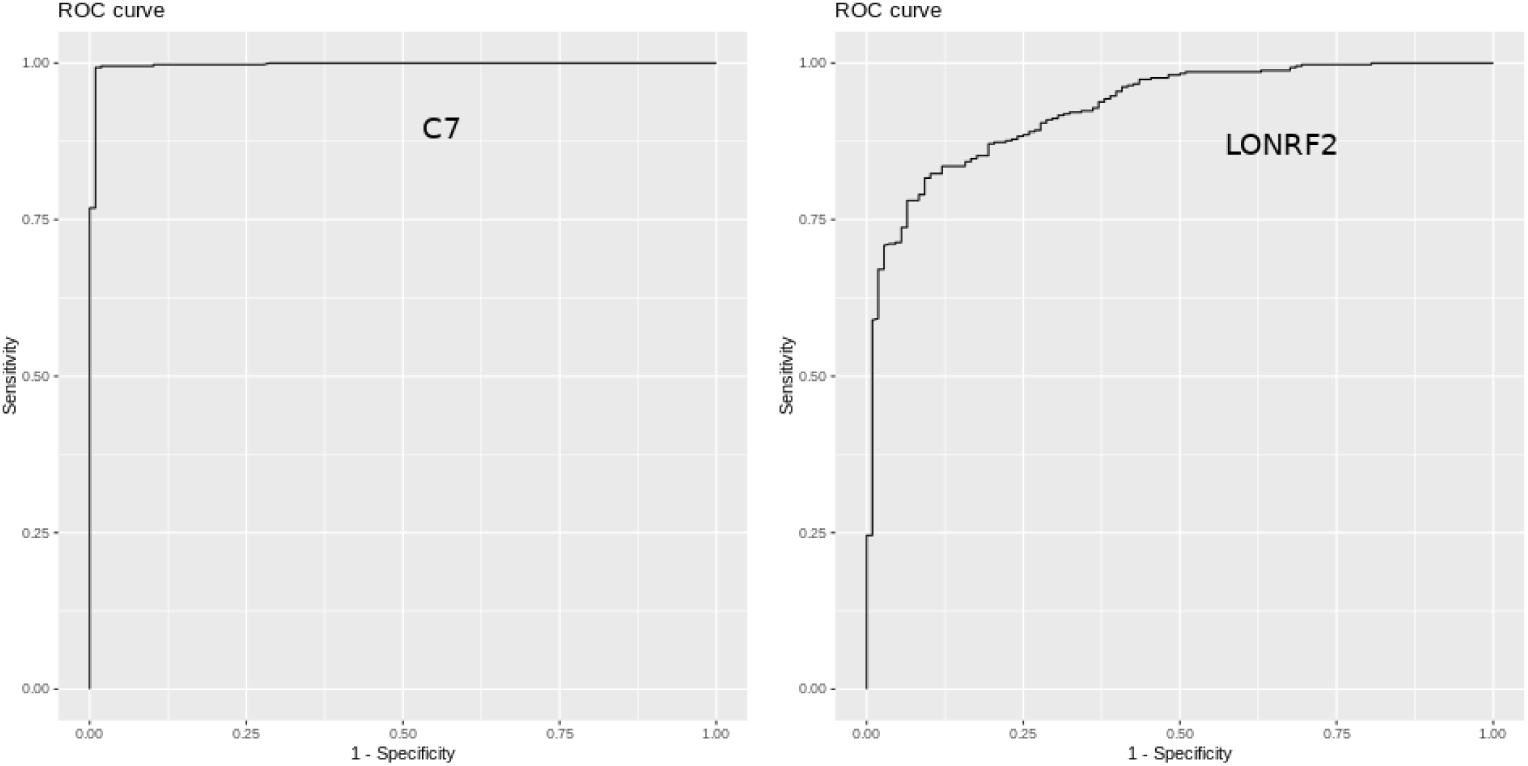
ROC curves for the OV singular biomarkers.

**Figure 21:**
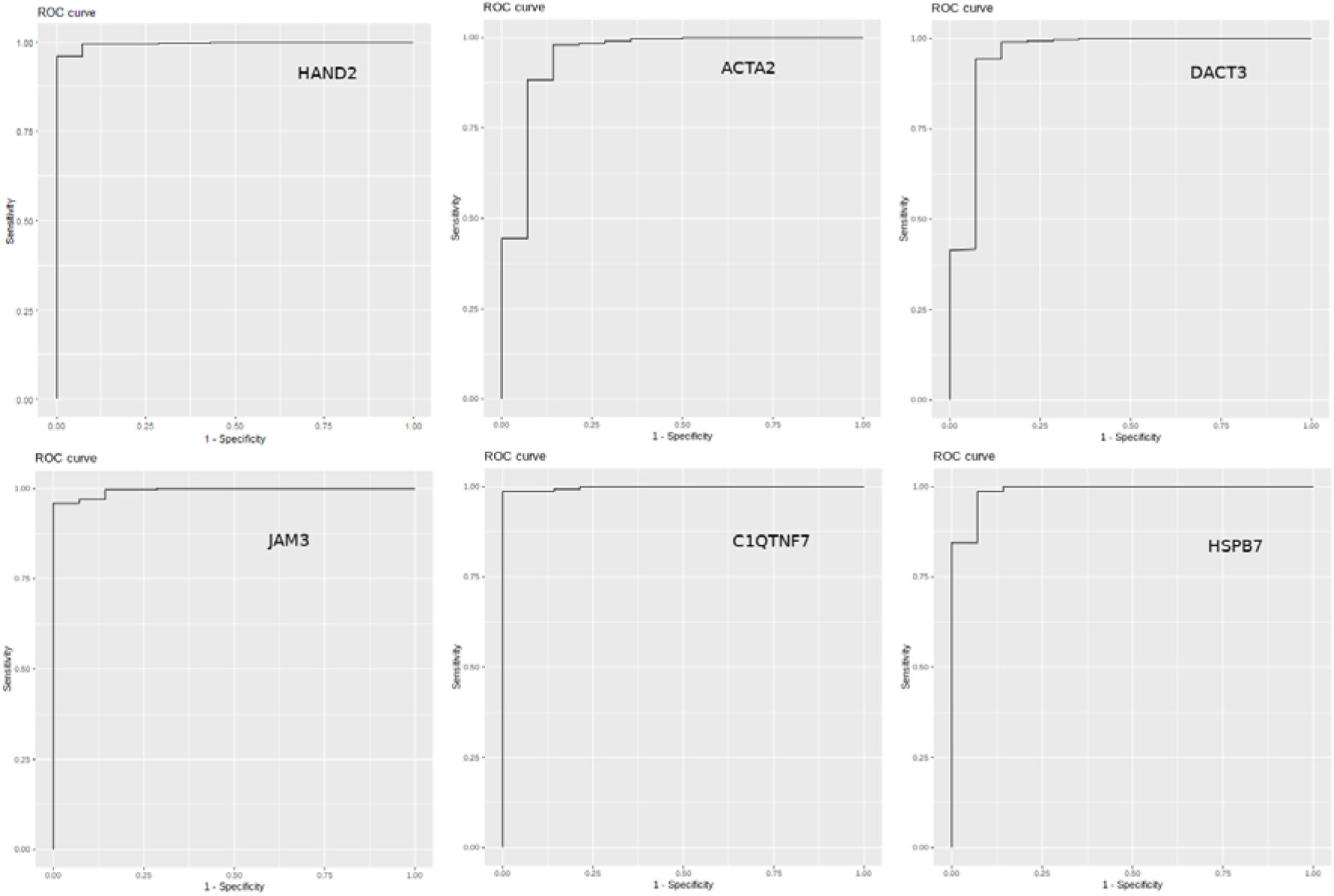
ROC curves for the CESC singular biomarkers.

**Table 4:**
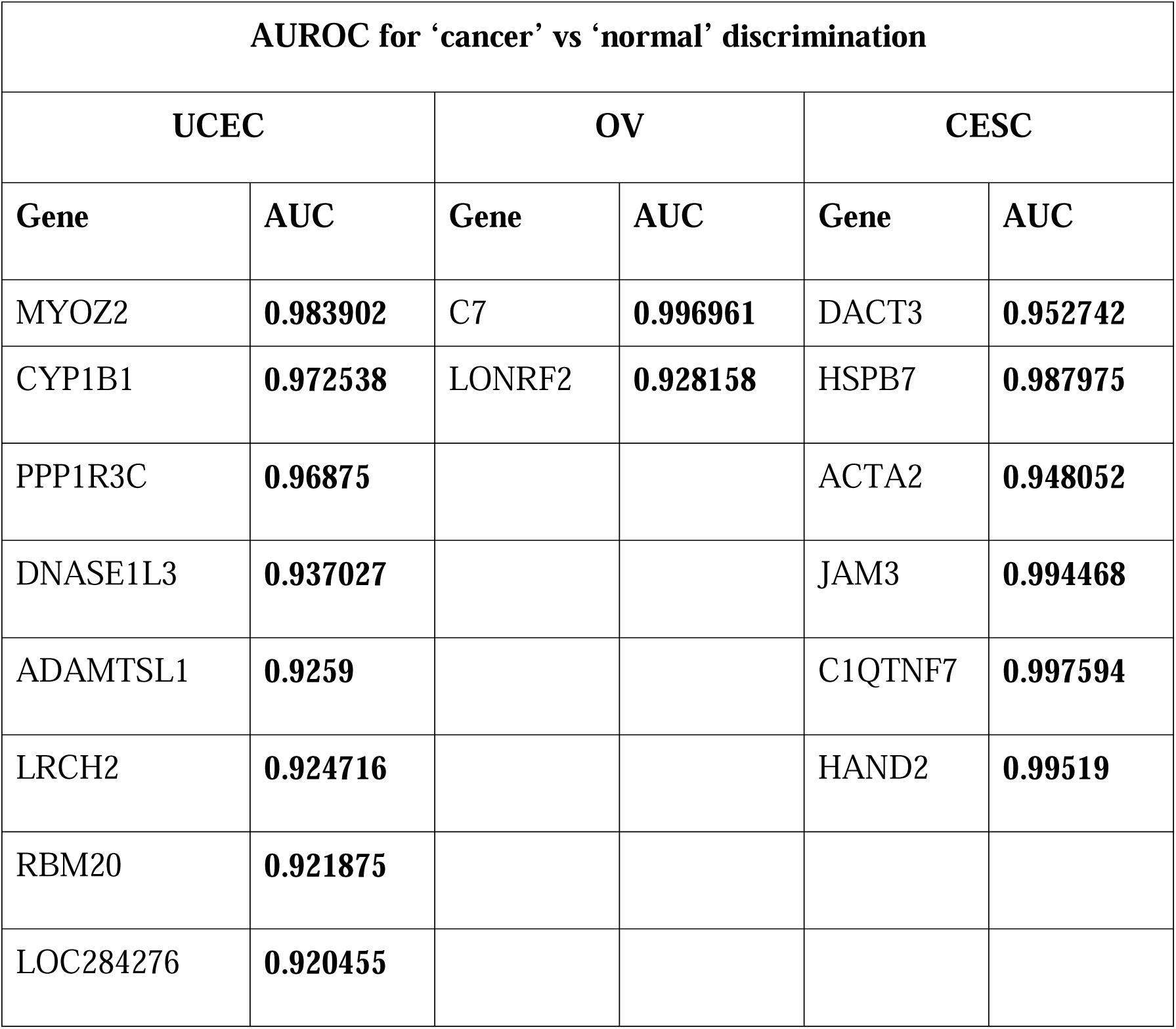
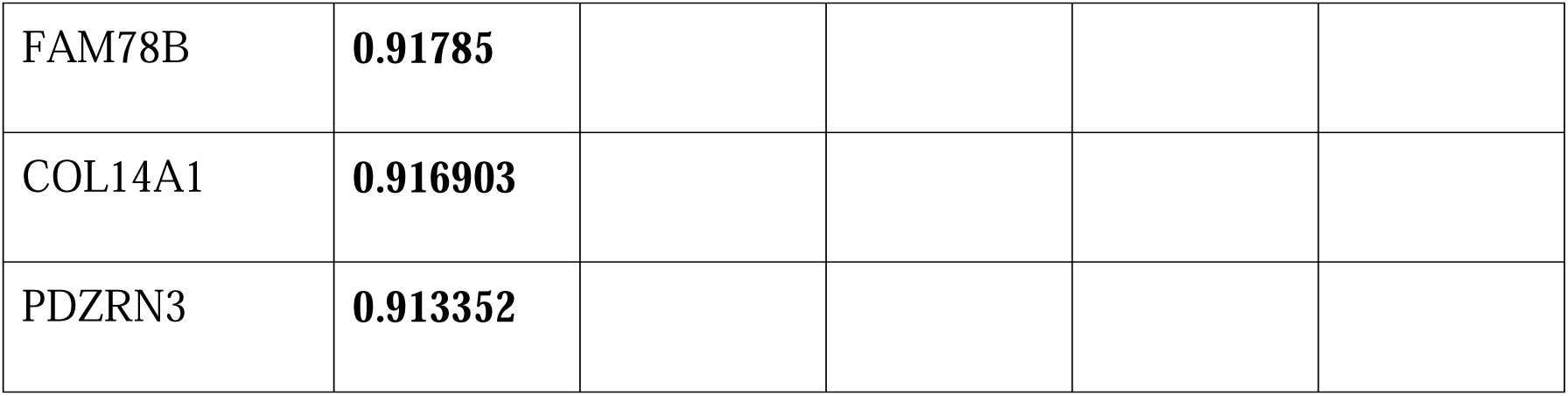
AUROC of individual consensus biomarkers for classifying cancer vs control for the cancers of interest. Only the singular biomarkers (AUROC > 0.9) are shown.

| **AUROC for ‘cancer’ vs ‘normal’ discrimination** |  |  |  |  |  |
| --- | --- | --- | --- | --- | --- |
| **UCEC** |  | **OV** |  | **CESC** |  |
| **Gene** | **AUC** | **Gene** | **AUC** | **Gene** | **AUC** |
| MYOZ2 | **0.983902** | C7 | **0.996961** | DACT3 | **0.952742** |
| CYP1B1 | **0.972538** | LONRF2 | **0.928158** | HSPB7 | **0.987975** |
| PPP1R3C | **0.96875** |  |  | ACTA2 | **0.948052** |
| DNASE1L3 | **0.937027** |  |  | JAM3 | **0.994468** |
| ADAMTSL1 | **0.9259** |  |  | C1QTNF7 | **0.997594** |
| LRCH2 | **0.924716** |  |  | HAND2 | **0.99519** |
| RBM20 | **0.921875** |  |  |  |  |
| LOC284276 | **0.920455** |  |  |  |  |
| FAM78B | **0.91785** |  |  |  |  |
| COL14A1 | **0.916903** |  |  |  |  |
| PDZRN3 | **0.913352** |  |  |  |  |

## CONCLUSIONS

The integration of multi-omics data yields a comprehensive investigation of the molecular basis of cancer pathology. Here, we have developed such a workflow and applied it to gynecological cancers. Our analysis of the three major gynecological cancers, namely cervical, ovarian and endometrial cancer, has identified stage-salient genes, consensus markers from multi-omics analysis and ultimately candidate singular biomarkers for each cancer. Such singular biomarkers include MYOZ2, CYP1B1, PPP1R3C, DNASE1L3, ADAMTSL1, LRCH2, RBM20, LOC284276, FAM78B, COL14A1, and PDZRN3 for endometrial cancer; C7 and LONRF2 for ovarian cancer; and HAND2, C1QTNF7, JAM3 HSPB7, ACTA2 and DACT7 for cervical cancer. The computational identification of molecular subtypes of each cancer was implemented and a proof-of-principle for a clustering-based screening model of endometrial cancer based on factor analysis of expression data has been demonstrated. It is hoped that these results would pave the way for early-stage detection models of gynecological cancers and their effective individualized treatment.

## Data Availability

All data produced in the present work are contained in the manuscript

https://portal.gdc.cancer.gov/

http://firebrowse.org

